# Nowcasting cases and trends during the measles 2023/24 outbreak in England

**DOI:** 10.1101/2025.04.08.25325363

**Authors:** Maria L Tang, Ian S McFarlane, Christopher E Overton, Erjola Hani, Vanessa Saliba, Gareth J Hughes, Paul Crook, Thomas Ward, Jonathon Mellor

## Abstract

**Background:** From 2023 to 2024, England had the largest measles outbreak in over a decade. Lags from suspected cases’ symptom onset to the availability of test results mean laboratory-confirmed case data are inherently retrospective rather than real-time. Reporting lags vary by measles prevalence and whether testing was for diagnostic or surveillance purposes. Nowcasting models predict future backfilling of reported cases and can estimate recent trends.

**Methods:** We developed a generalised additive model framework accounting for reporting delays, location, and day-of-week effects in line-list data by onset date. The model was re-fit weekly providing real-time nowcasts and directional trends for national and regional users. Retrospectively, we tested alternative specifications to optimise structure and confirm predictive performance, evaluating with log weighted interval score (WIS) and ranked probability score (RPS).

**Results:** For case count estimates, the operational and retrospective models outperformed the baseline model, with a lower average daily national log WIS by 42% and 41%, respectively, and similar regional improvements. For four-week trend direction, the operational and retrospective models provided better national estimates than the baseline with an average RPS lower by 69% and 6% respectively. Regionally, they also outperformed the baseline model in London, but the baseline model offered better performance for the smoother single-peak West Midlands epidemic. An alternative model indexed by report date instead sometimes outperformed the other nowcasting models for trend direction but also lagged changes in trend.

**Interpretation:** Our work highlights the utility for real-time nowcasting models during outbreaks to inform fast-evolving trends, and the need for early access of accurate reporting delay data to facilitate effective modelling.

## Introduction

In January 2024, a national incident was declared for measles in England by the UK Health Security Agency (UKHSA) after a rapid increase in cases in the West Midlands region [1]. This was followed by a large outbreak in London and smaller clusters in other regions, becoming the largest measles outbreak in England in over a decade. Measles is a highly contagious disease spread through airborne transmission [2]. Patients present with a prodrome of cold-like symptoms, conjunctivitis, and fever, followed by a rash. Measles can be serious and lead to complications like ear infection, chest infection and encephalitis, with infants, pregnant women and immunocompromised adults being the most vulnerable [2]. Measles was declared eliminated in the UK in 2023 through sustained high uptake of the combined Measles-Mumps-Rubella vaccine [3], however uptake of the routine childhood immunisation programme declined over the last decade [4], increasing the pool of susceptible children and therefore leading to larger importation-driven outbreaks.

The 2023/24 measles outbreak in England was characterised by delayed reporting of cases, resulting in incomplete real-time data, making interpretating recent trends challenging. These inherent delays between symptom onset date and case reporting encompass healthcare-seeking and laboratory-testing time, varying geographically and by test type. UKHSA reporting of laboratory-confirmed measles cases by symptom onset date includes a caveat on reporting delays to aid interpretation of the most recent four weeks. To address these limitations, nowcasting models provide estimates of the most recent data, accounting for reporting delays.

To account for reporting delays, nowcasting methods have been applied widely to disease outbreaks [5] [6] [7] [8] [9]. A flexible generalised additive model (GAM) was developed to nowcast the 2022 mpox outbreak in England, leveraging individual-level patient and reporting delay data to estimate real-time case counts [7]. This framework was further applied successfully to address reporting delays in seasonal norovirus surges in England [8], highlighting its adaptability to modelling different pathogens. Despite the utility of nowcasting for infectious disease surveillance, there has been limited application to measles, the most notable example being the retrospective study nowcasting of a measles outbreak in the Netherlands in 2013-2014 [9].

Public health decision makers are interested in epidemic trends in addition to case count estimates. Trends are communicated through estimated quantities like growth rates [7] [10] and the effective reproduction number *R*_*t*_ [10] [11], inferred from modelling. Probabilistic ordinal trend categorisation, such as a signal increasing, decreasing, or being stable, is more commonly applied in economics and finance [12] [13]. Ordinal categories provide quick and useful indications of an epidemic’s status for both technical and non-technical decision makers and so, here, was preferred by stakeholders to more complex indications of trend.

This paper presents a nowcasting model developed during the 2023/24 measles outbreak in England providing near real-time weekly estimates of case counts and epidemic trend direction. Building on previous nowcasting GAMs [7] [8] and based on the needs of stakeholders, our model estimated regional and national case counts, accounting for differences in reporting delay according to region and test type, with ordinal trend categorisation. Given the time constraints for model development during the outbreak, we retrospectively assess the operational model’s performance against a model optimised over the whole outbreak and a baseline model. Our work highlights the value of nowcasting during a large infectious disease outbreak to improve real-time situational awareness of evolving epidemic trends and support timely outbreak response and decision making.

## Methods

### Data

#### Data flow

UKHSA Health Protection teams send an oral fluid kit to every suspected measles case for surveillance purposes, in accordance with World Health Organisation measles elimination requirements [3]. Samples are then sent to the National Reference Laboratory for testing. Cases that attend hospital are also likely to undergo diagnostic testing with samples first sent to local National Health Service or UKHSA laboratories. This is the faster route to obtain results as it is intended to inform case or contact management. Locally-confirmed cases should also have their samples forwarded onto the reference laboratory for confirmatory testing. The time taken to process a sample in the reference laboratory is more sensitive to changes in demand during outbreaks. Cases with a positive laboratory test become “confirmed” and are added to the outbreak line list, although a small subset may be temporarily set aside for further review (Supplementary Section 1). In this analysis, the effect of data quarantine was not considered due to data constraints.

#### Line list data

We used the line list of laboratory-confirmed measles cases extracted on 8 October 2024 from UKHSA databases. Throughout the outbreak, the line list was updated on Tuesday each week with new data up to the previous Monday evening. Cases in the line list were confirmed by either a local laboratory test, reference laboratory test or a local laboratory test followed by a confirmatory reference laboratory test (Supplementary Table 2).

Cases were included in the model training data if they had a symptom onset date (0% missing) and at least one of local laboratory test received date or reference laboratory test report date (3.5% missing) (Supplementary Table 3). Confirmed cases had one or both test dates, so we defined the *earliest report date* as the earliest of the two and used this as the single report date in the models. This represents the estimated earliest date that a confirmed case is added to the measles line list. We further excluded cases from the model training data if the reported symptom onset date was later than the earliest report date, i.e., symptoms were reported to have started after the positive test, true for 0.7% of cases.

Date definitions

- **Symptom onset date:** date when the case first presented with symptoms.
- **Local laboratory test received date**: date the local laboratory test result was received in the UKHSA database.
- **Reference laboratory test report date:** date the reference laboratory test result was reported to the patient’s general practitioner surgery, a proxy for when the result was received in the UKHSA database.
- **Earliest report date:** the earliest of the two (local laboratory test received date and reference laboratory test report date), a proxy for the date that a confirmed case is added to the measles line list. Ties are excluded (5 cases in total). This is the single report date used in the models.

### Model

Our aim was to produce a nowcasting model to estimate the number of confirmed measles cases in recent weeks, including the cases that had yet to be reported, referred to as a nowcast. To support management of infectious disease outbreaks, both national and regional case count estimates for regions with ongoing outbreaks were required, as well as estimated recent trend. These trends help assess whether the current trajectory of the epidemic is increasing, stable or decreasing, based on agreed thresholds. The first nowcast product was delivered on 3 July 2024, after development, validation and user testing of the model and product. The product was delivered weekly initially, before frequency dropped to biweekly and monthly, with the last nowcast product delivered on 23 October 2024, following de-escalation of the incident.

During the outbreak, the models were sequentially re-fit each week on the updated training data to provide new estimates for the most recent weeks. Line lists were updated each Monday evening, with the training data truncated to Sundays to remove the incomplete final day and align with wider reporting. In this paper, we present the predictions between the weeks ending 3 July 2024 and 8 September 2024 from the operational model, covering the period of weekly deployment during the outbreak. For all other models, we considered the model performance from near the beginning of the outbreak, from weeks ending 3 December 2023 until 8 September 2024.

We define terms to clearly express the different time periods associated with the prediction and model fitting processes:

- **Final prediction date**: the reference date for separate model fits – the date up to which a model is fitted using all available data and the last date for case count predictions.
- **Nowcast horizon**: the 4-week period over which case count predictions are made, with days and weeks indexed as non-positive numbers, where day 0 is the final prediction date and day - *x* is *x* days prior.

To estimate case counts using the operational and retrospective models detailed in the next sections, we decompose *N*_*t*_, the number of cases with symptom onset date *t*, by their reporting delay:

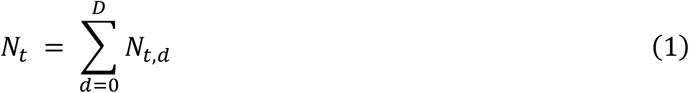

where *N*_*t,d*_ is the number of cases with symptom onset date *t* = − *T* + 1, …, 0 reported after *d* days delay, *d* is the number of days between symptom onset and earliest report date, and *T* is the model training length.. We only fit to recent delay data from the last *D* ≤ *T* days. As the data are right-truncated, this sum consists of case counts known at time 0, *N*_*t,d*_ for *t* + *d* ≤ 0, and case counts that are unknown and yet to be reported, *N*_*t,d*_ for *t* + *d* > 0. Hence, to obtain an estimate for the whole *N*_*t*_, we only need to estimate the unknown *N*_*t,d*_ and sum them with the known *N*_*t,d*_.

To generate region-specific nowcasts, the nowcasting models incorporate region structure. Regions that did not experience large outbreaks (North West, North East, East of England, East Midlands, South East, and South West) were aggregated into an “Other” category to consolidate low counts. Since the operational model was deployed mid-outbreak, after the West Midlands outbreak, it used only two regions: London and “Other + West Midlands”. The retrospective models cover the whole outbreak, so include three regions: West Midlands, London and “Other”. For evaluation, only national, London and West Midlands predictions are considered, because the “Other” region was not relevant to stakeholders. National predictions were aggregated from all regions:

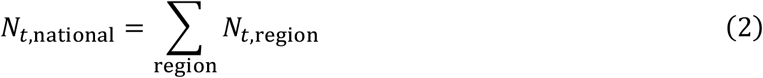

To generate a nowcast, we use generalised additive models (GAMs), a flexible framework for modelling smooth time-varying components, based on previous work nowcasting mpox [7] and norovirus outbreaks [8]. For real-time nowcasting in an incident, GAMs were easy to adapt from existing models to deploy and run quickly each week. We model the unknown integer count data using a negative binomial distribution to account for overdispersion, and a log link function to model the exponential epidemic process. The models described below follow a general form modelling *N*_*t,d*,subgroup_the number of cases with symptom onset date *t* reported after *d* days delay in a particular subgroup (region, test type):

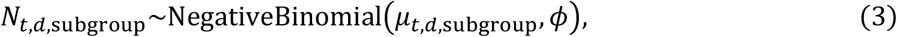

with dispersion parameter *ϕ* and mean *μ*_*t,d*,subgroup_, where

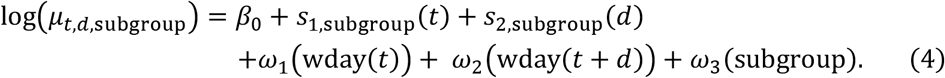

Here we have

- *β*_0_ is a constant,
- *s*_1,subgroup_(*t*) is a subgroup-specific smooth over symptom onset date,
- *s*_2,subgroup_(*d*) is a subgroup-specific smooth over reporting delay,
- *ω*_1_(wday(*t*)) and *ω*_2_(wday(*t* + *d*)) are day-of-week random intercepts for symptom onset date and earliest report date, respectively,
- *ω*_3_(subgroup) is a subgroup random intercept.

All models were fitted in R using the *gam* function from the *mgcv* package [14]. 1000 posterior samples were generated using the *gratia* package [15]. Daily regional samples were aggregated to daily national, weekly regional and weekly national levels. Prediction intervals are extracted from the posterior samples at each of these levels.

### Operational nowcasting model

Our model builds on previous nowcasting models [7] [8], incorporating additional disaggregation of reporting delay by region and test laboratory type to account for differences seen in reporting delay between test types (Supplementary Table 3) and between London and other regions for the same test laboratory type (Table 1). Hence, we model *N*_*t,d*_ additionally stratified by test type and region as *N*_*t,d*,test type,region_:

**Table 1:**
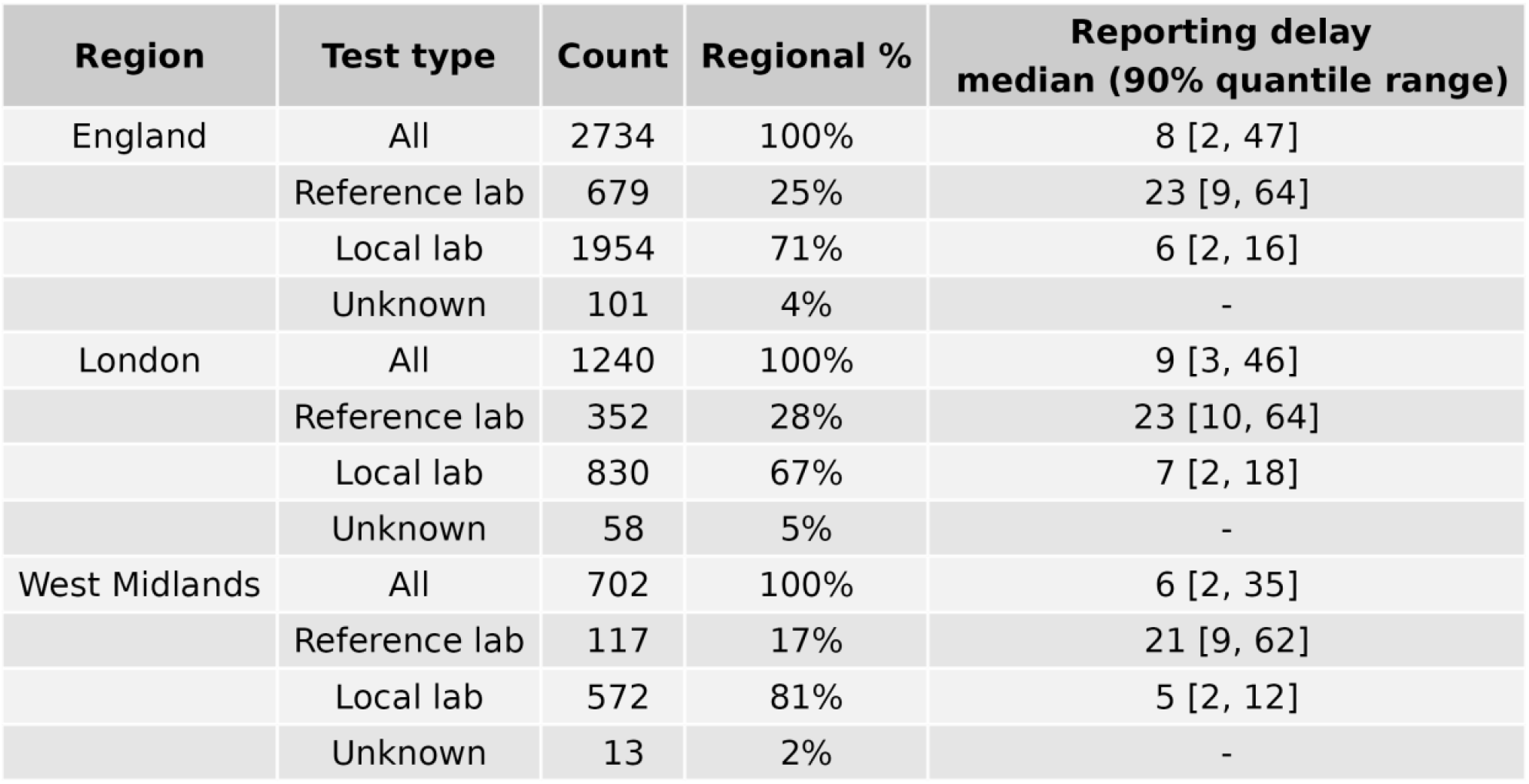
Count of cases by region, earliest test type and their associated reporting delay (days since symptom onset). Cases with “unknown” test type either have no reference laboratory and no local laboratory test report date or have equivalent reference laboratory and local laboratory test report dates (5 cases).

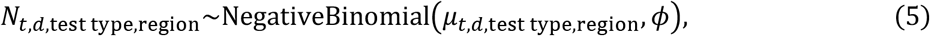

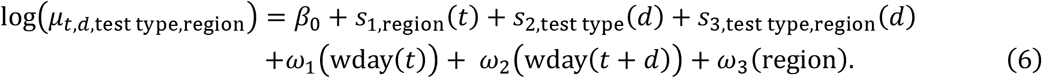

*s*_3,test type,region_(*d*) is a smooth over reporting delay that gives the deviations from the test type smooths by region. The model used a training length of 105 days, maximum delay of 42 days and thin plate regression splines with the default basis size of 10, based on initial exploration. Model structures and hyperparameters were compared at the time with limited data, but full exploration and tuning were not possible due to time constraints in deploying a timely product.

### Retrospective nowcasting model

As full tuning was not completed for the operational model, we retrospectively explore different model structures and hyperparameter tuning by evaluating predictions over the extended outbreak period to find an overall optimal model structure (Supplementary Section 2). The resulting model is similar to the operational model but does not include test type:

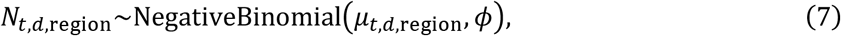

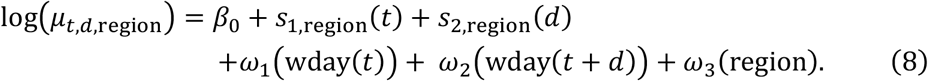

Model tuning (Supplementary Section 2) determined hyperparameter values and model structure. The model uses a training length of 105 days, maximum delay of 42 days and thin plate regression splines with a basis size of 15 for *s*_1,region_(*t*) and 6 for *s*_2,region_(*d*).

### Baseline model

For a baseline model, we aim to simulate the trend that a decision maker viewing the epidemic curve would assume in the absence of a nowcast. Since reporting delays are mostly less than 14 days (75%, Supplementary Figure 1), the baseline model assumes that case counts earlier than 14 days ago will remain the same as the currently known data, with all prediction intervals equal.

Regional case counts are thus given by

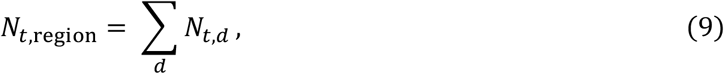

for *t* = −27, …, −14. For the following 14 days, an extrapolated trend is assumed. We fit a GAM to the data excluding the most recent 14 days and forecast the next 14 days assuming a constant growth rate, following short-term forecasting methods [16]. Regional case counts are modelled directly as *N*_*t*,region_:

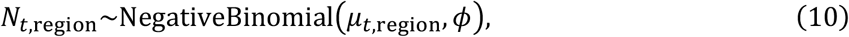

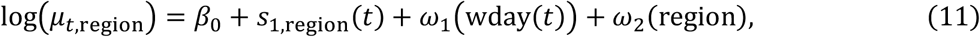

for *t* = −13, …, 0. Hyperparameters chosen from model tuning (Supplementary Section 2) were a training length of 147 days and cubic regression smooths for *s*_1,region_ with 7 knots.

### Report date model (for growth rate and trends)

As an additional model for comparing growth rate and trends, we consider a simple model defined by earliest report date rather than symptom onset date. This approach aligns with users who may view the data by report date to work with a complete time series, which can then be used to assess recent trends. The model becomes

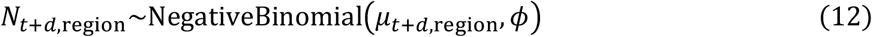

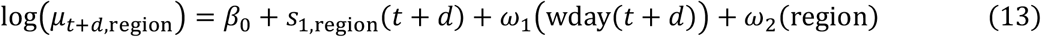

where *t* + *d* is the earliest report date. The case count estimates by report date are not used directly, but instead used only for calculating trends and growth rates. Hyperparameters chosen from model tuning (Supplementary Section 2) were a training length of 105 days, and Gaussian process smooths for *s*_1,region_, with a linear trend and power exponential covariance function with power 1.5, and range parameter 15.

### Trend categorisation

Alongside case count estimates, we provided decision makers with estimated national and regional (London and the West Midlands) trend directions (increase, stable, or decrease) and their associated probabilities. We presented a range of trend direction thresholds and time periods to stakeholders, and agreed upon ±0.1 and comparing the most recent four weeks to the previous four weeks, capturing changes that were most meaningful to stakeholders based on operational needs. We also explored shorter time periods for more reactive trend changes.

To calculate the most recent *n*-week trend, we first sum estimated case counts for the most recent *n* weeks and subtract it from the sum of estimated case counts from the previous *n* weeks. For model prediction sample *i*, we calculate the change in case counts in the most recent *n* weeks for each region, Δ_*n*,region,*i*_, as

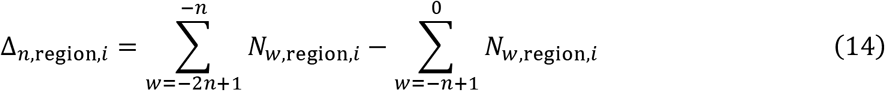

where *N*_*w*,region,*i*_ is the estimated regional case count for week *w* up to the final prediction week 0 for model prediction sample *i*. The trend is then assigned based on the thresholds:

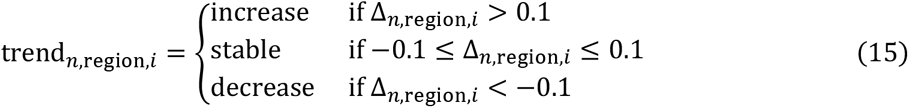

Probabilities are calculated as the proportion of samples *i* falling within each trend category. Trends were calculated separately nationally and for regions with ongoing outbreaks. Equations 13 and 14 are similarly applied to national estimated counts.

In addition, growth rates were explored for evaluating trends (Supplementary Section 3).

## Evaluation

To tune model hyperparameters and compare models, we use the *scoringutils* package [17]. For case counts, we only score the last four weeks of predictions from each model fit, as this period is of the most use (88% of delays are within four weeks - Supplementary Figure 1) and is the period considered incomplete in official reports. We primarily score case count estimates by the weighted interval score (WIS) on a log scale. The WIS is a proper score that balances calibration (how well the predicted distribution matches the outcomes) and sharpness (width of prediction intervals), with lower values indicating better performance [18]. Transforming to a log scale can better measure the performance of predicting the exponential growth rate of an epidemic [19]. We calculate the WIS using the median and 50% and 90% prediction intervals. Additionally, we assess model bias, which shows whether predictions usually overestimate (bias > 0) or underestimate (bias < 0), and the 90% coverage, giving the proportion of true values falling within the 90% prediction intervals. Case count predictions are scored at the daily and weekly temporal resolutions, as well as both national and regional geographic resolutions, focusing on regions that experienced an outbreak (London and the West Midlands). Additionally, we consider scores by day and week across the nowcast horizon to assess differences in performance throughout the four-week period of predictions.

For trend categorisation, we score using the ranked probability score (RPS), a proper scoring rule for ordinal probabilistic predictions that is 0 for a perfect score and positive otherwise. The RPS is “sensitive to distance”, meaning it penalises predictions more if they are further from the observation according to an ordering of categories (in our case: decrease, stable, increase) [20].

To compare models against the baseline model, we calculate skill scores using WIS and RPS, which quantify by how much a model improves predictions compared to the baseline model, according to the chosen scoring rule:

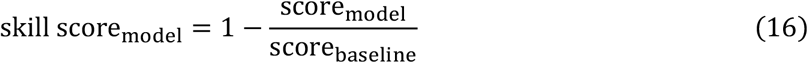

## Results

### Descriptive

The 2023/24 measles outbreak in England was mostly influenced by the initial West Midlands outbreak until January, when cases emerged in other regions, causing a plateau of high case numbers before a decline from August (Figure 1). Regionally, the West Midlands outbreak peaked sharply in early January, followed by an outbreak in London with a broader peak (Supplementary Figure 2). Other regions experienced smaller outbreaks. When analysed by report date, the national epidemic curve looks similar (Figure 1), but with the West Midlands peak lagged by one week, and the London plateau also lagged (Supplementary Figure 3).

**Figure 1:**
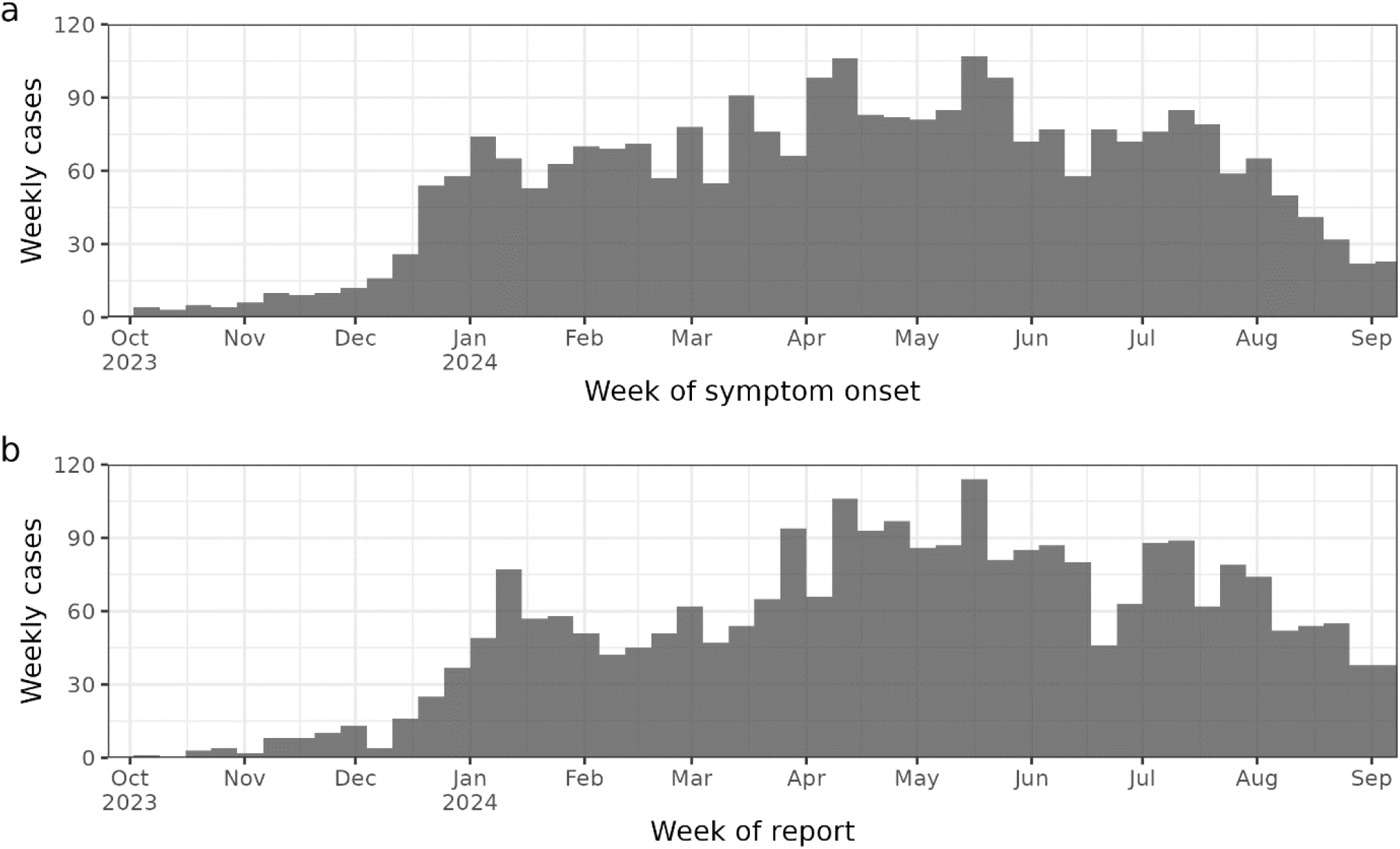
Count of confirmed measles cases by week according to (a) symptom onset and (b) earliest report date.

Reporting delay, the difference between symptom onset and earliest report date, varied by test type and region. Reference laboratory tests had longer and more variable delays than local laboratory tests (Table 1), with both showing long-tailed distributions (Supplementary Figure 4). Reporting delays varied over time, with mid-January to mid-February seeing a spike in reference laboratory delays and longer delay distribution tails (Supplementary Figure 5), coinciding with the peak in West Midlands cases. Regionally, London experienced longer median delays for both test types. The Reference Laboratory undertook a higher proportion of all overall tests in London which likely contributed to longer delays overall (Table 1) and during periods of high measles activity mid-January to late-March (Supplementary Figure 6).

### Model predictions

Figure 2 and Supplementary Figures 7 to 21 show model predictions over all weeks at daily national, daily regional, weekly national and weekly regional scales. The narrow prediction intervals in weeks −4 and −3 of the nowcast horizon for all models result in substantial errors when there are more revisions than usual, such as in the model fit ending 25 February 2024 (Figure 2). The baseline model’s forecast in weeks −1 and 0 of the nowcast horizon can perform badly when the trend changes, particularly at turning points (e.g. Supplementary Figure 13).

**Figure 2:**
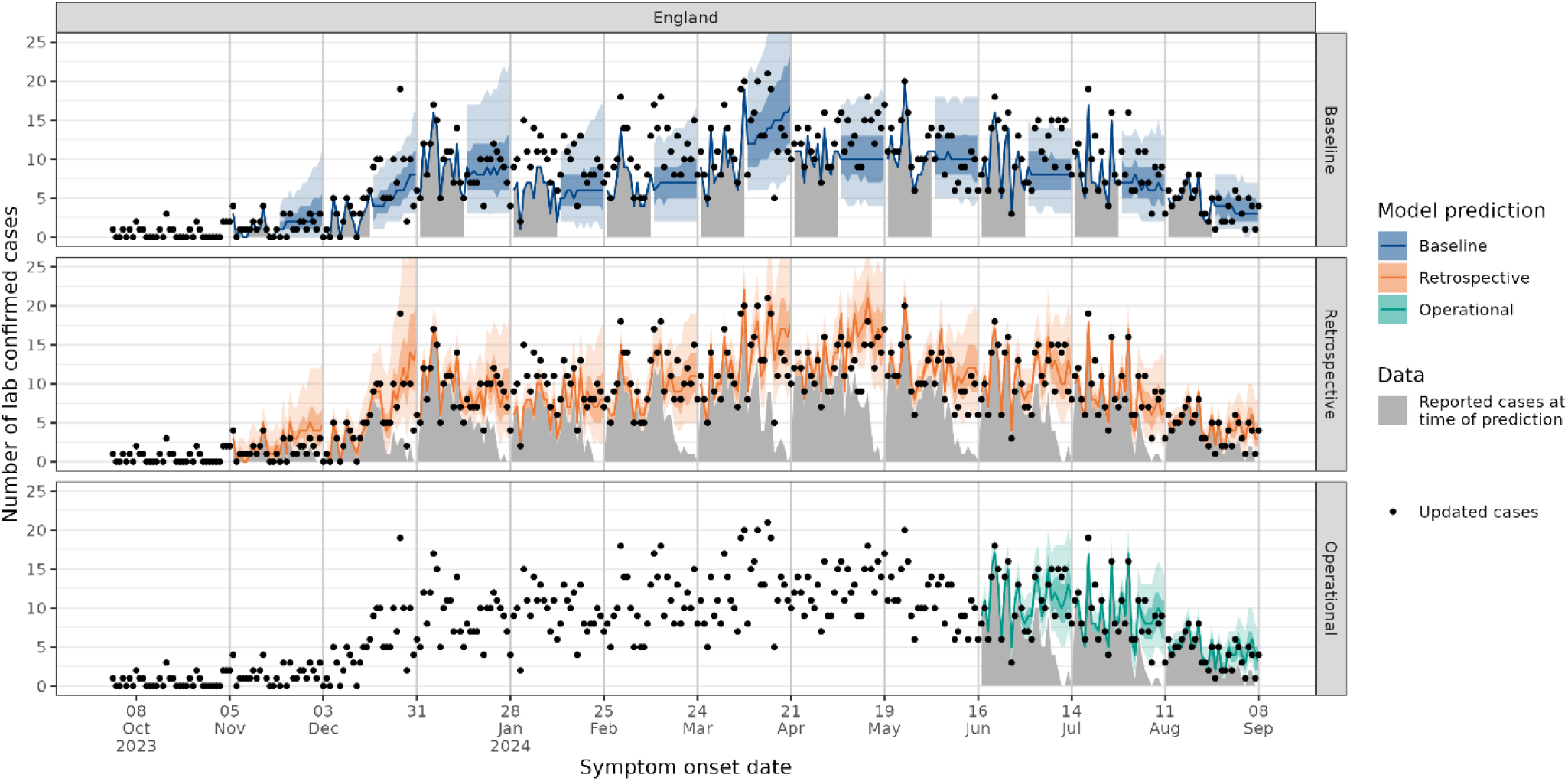
Daily national predictions (median, 50% and 90% prediction intervals) for models re-fitted on new available data each week. Every fourth model fit is shown for clarity, with the first fitted on data up to 3 December 2023. The other model fits are given in Supplementary Figure 7 to 9. The baseline model uses initial data (shaded in grey) only for predictions in weeks −4 and −3 of each nowcast horizon. The operational model is shown only for the period it was used during the outbreak.

On the daily scale, the operational and retrospective nowcasting models perform better than the baseline model, with respective smaller log WISs by 42% and 41% nationally, 41% and 39% in London and 37% for the retrospective model in the West Midlands (Supplementary Figure 22). The operational and retrospective models also outperform the baseline model for almost all model fits across the outbreak (Figure 3). The operational model scores similarly to the retrospective model in the weeks when the operational model was used. Aggregated weekly, the nowcasting models also perform better than the baseline, although scores are more similar, with the baseline performing better in several more weeks (Supplementary Figure 23). Across the nowcast horizon, the operational model slightly outperforms the retrospective model, and both outperform the baseline (Supplementary Figure 24, 25). Log WIS scores generally worsen for all models closer to day 0, but also converge, suggesting that these predictions are difficult for all models. The retrospective and baseline model perform more similarly between days −14 and −27 in the West Midlands than in London and England, when the baseline’s assumption of no backfilling during this period performs well. On a weekly level, the retrospective and operational models show an improvement in coverage in week 0 as wider prediction intervals in this week capture more of the observations (Supplementary Figure 251).

**Figure 3:**
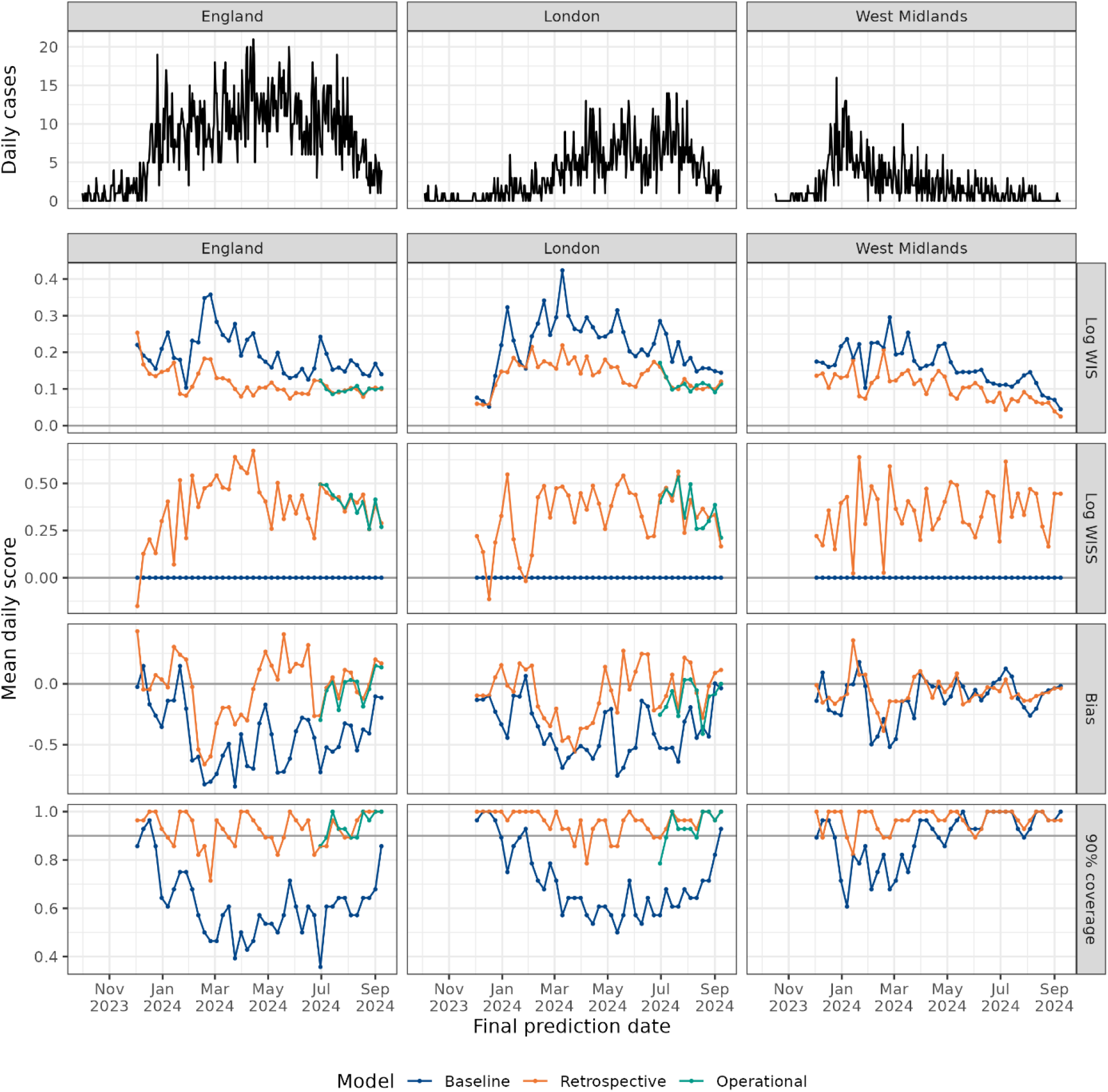
Daily cases over time in England, London and the West Midlands (top pane), and mean daily scores by prediction end date and model: log weighted interval score (WIS), log weighted interval skill score (WISS), bias and 90% coverage. The operational model is shown only for the period it was used during the outbreak.

### Trend categorisation

For four-week trends, all models outperform the baseline model nationally, with the operational, retrospective and report date models’ average RPS lower by 69%, 6% and 21% respectively (Supplementary Figure 26). All models capture the initial national increase well but are worse at predicting the following stable periods (Figure 4b). The baseline model performs worst around trend transitions, but performance usually improves if that trend is sustained, and the model extrapolation adjusts to the new trend (peaks in baseline RPS, Figure 4c). For the single-peak West Midlands outbreak, which features a period of increase followed by a mostly decreasing trend, the baseline model outperforms the retrospective and report date models, capturing the decrease more consistently. However, for the multi-peak London outbreak, the operational and retrospective models show a greater improvement over both the baseline and report date models.

**Figure 4:**
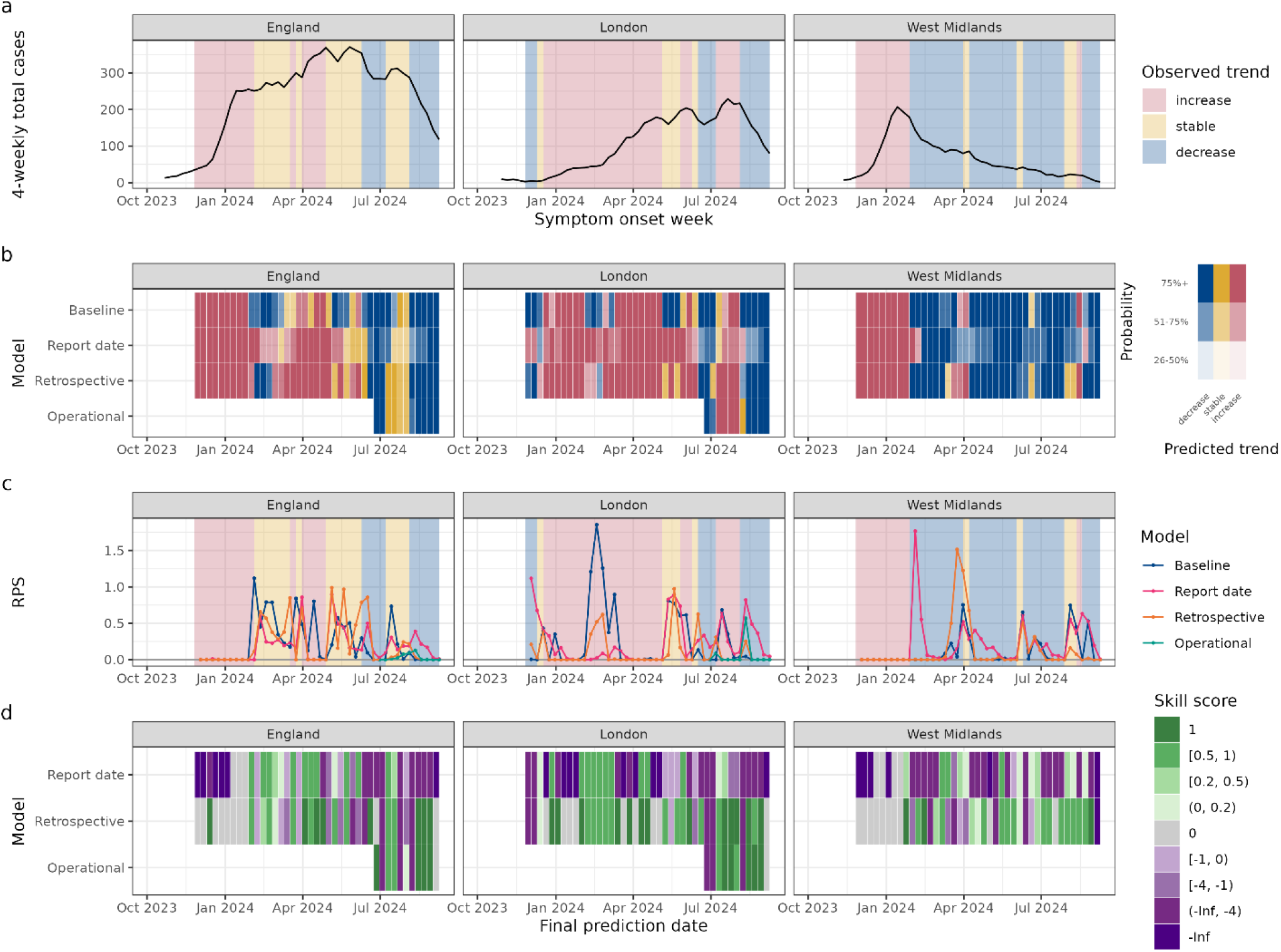
(a) Four-week rolling sum of cases. (b) Predicted most likely four-week trend. (c) Ranked probability score (RPS) – lower is better. (d) Ranked probability skill score: green (better than baseline), grey (same), purple (worse). The operational model is shown only for the period it was used during the outbreak.

The report date model performs better nationally than the retrospective model, but worse in London and the West Midlands. The report date model is seen to sometimes react slower to trend changes, most notably at the West Midlands peak, where its predicted change in trend lags by two weeks. In London, the retrospective and operational models are similar and outperform the report date model, capturing local changes better. Nationally, the operational model outperforms all other models over the weeks it is in use. In addition to the lag at detecting changes in the trend category, the growth rate calculated from the report date model also lags the growth rate calculated from the nowcasting models (Supplementary Section 3).

For one-week trends, models perform more similarly overall (Supplementary Figure 27), as all models struggle to predict the highly variable short-term trends (Supplementary Figure 28).

## Discussion

This study presents the real-time implementation of a nowcasting model applied to a measles outbreak in England, estimating case counts and trends. Nowcasting becomes more challenging over a longer period, so this outbreak was interesting to model due to its long reporting delays (needing revisions to the last four weeks of data), and the variation in reporting delay by region and testing laboratory type. Our model flexibility ensured rapid development and deployment of a weekly output providing timely insights during the outbreak. This nowcasting model framework was effective not only on the weeks the operational model was deployed, but also when applied retrospectively to the whole outbreak. Our approach outperformed a baseline model that forecasted a trend from the prior two weeks, offering more reliable case count estimates, especially around epidemic peaks.

Reporting delay varied between regions and test types, with both factors incorporated into the operational nowcasting model’s delay structure. Retrospectively, a simpler delay structure by region only was sufficient to capture delay variations over the whole outbreak and were important in estimating regional case counts, outperforming other alternative model structures explored. Too many breakdowns, particularly when related, can add uncertainty, especially early in an outbreak with few cases. We saw that breakdowns were more important in estimating case counts regionally compared to nationally. The importance of different breakdowns may change over an outbreak, emphasising the importance for real-time monitoring and adjustments.

Reporting delay distributions also varied over time. Although our models use the most recent delay data, a limitation is assuming that this distribution continues, which cannot account for rapid changes. For some weeks, the retrospective nowcasting model performed badly when more tests than expected were reported in the earlier weeks that usually see few revisions. Factors such as changes in test volume and laboratory capacity can affect reporting delay, especially for reference laboratory tests which are processed centrally in a single national laboratory. Incorporating an explicitly time-varying delay distribution could improve model accuracy, but at the expense of computation time and complexity.

Our customers preferred ordinal trend categorisation to growth rates for understanding changing trends, although suitable thresholds need to be defined. Four-week trends were easier to predict than shorter, more variable trend periods. Overall, the operational and retrospective nowcasting models generally outperformed the baseline model for noisier signals, but the baseline method of trend extrapolation worked well for the West Midlands’ single-peak epidemic with clear growth and decline periods, which was also easier to predict by all models. The report date model performed well nationally, but as report date falls chronologically after symptom onset date, was sometimes slow to react to trend changes, with observed lags around the length of the average reporting delay.

These models only require individual-level symptom onset and report dates, a convenient simplicity given the limited data availability and accessibility, but with limitations. We use earliest report date as a proxy for when cases were added to the line list. This does not account for additional delays due to quarantine review or cases without a report date. Although these cases are a minority, recording line list inclusion dates could improve model accuracy. We later received these data, but weekly censoring and the need for over a month’s data for model training prevented its timely integration into the product. Early data access is especially important for outbreaks with long reporting delays to have enough data for model training, although without this at the start of an outbreak, a delay distribution could be assumed from historical data.

For the operational model, we balanced model simplicity, reducing both model development time during the outbreak and computational running time, and performance. Our estimates are based only on the partial case data and reporting delay distributions, and models do not explicitly account for external factors, such as changes in vaccination rates, behaviour, or testing practices. Some of these factors may nonetheless be indirectly reflected in changes to the partial data and delay distributions which in turn influence model estimates. However, as previously mentioned, the model is less reactive to sudden changes in these external factors.

Overall, we have presented several different nowcasting models for estimating case counts and trends that are quick to implement and develop for fast-paced incident response. These models depend on and emphasise the importance of relevant data collection, particularly an accurate report date, as early as possible during an outbreak, and investigating the categorical breakdowns that show the greatest variation in cases and delay distributions. Further, actively engaging stakeholders in UKHSA was vital in ensuring that nowcasting products are accessible and align with practical needs in incident response. By utilising nowcasting products early in outbreaks, public health agencies can improve situational awareness and respond more effectively to emerging health threats.

## Data Availability

Code for the paper is available at: https://github.com/maria-tang/measles-nowcast
Aggregate annual measles cases are provided on the gov.uk website https://www.gov.uk/government/publications/measles-epidemiology-2023. Individual-level data including reporting delays have not been shared due to patient identifiability.
UKHSA operates a robust governance process for applying to access protected data that considers:
- the benefits and risks of how the data will be used
- compliance with policy, regulatory and ethical obligations
- data minimisation
- how the confidentiality, integrity, and availability will be maintained
- retention, archival, and disposal requirements
- best practice for protecting data, including the application of "privacy by design and by default", emerging privacy conserving technologies and contractual controls.
Access to protected data is always strictly controlled using legally binding data sharing contracts.
UKHSA welcomes data applications from organisations looking to use protected data for public health purposes.
To request an application pack or discuss a request for UKHSA data you would like to submit, contact.

## Data Availability

Aggregate annual measles cases are provided on the gov.uk website https://www.gov.uk/government/publications/measles-epidemiology-2023. Individual-level data including reporting delays have not been shared due to patient identifiability.
UKHSA operates a robust governance process for applying to access protected data that considers:
- the benefits and risks of how the data will be used
- compliance with policy, regulatory and ethical obligations
- data minimisation
- how the confidentiality, integrity, and availability will be maintained
- retention, archival, and disposal requirements
- best practice for protecting data, including the application of "privacy by design and by default", emerging privacy conserving technologies and contractual controls.
Access to protected data is always strictly controlled using legally binding data sharing contracts.
UKHSA welcomes data applications from organisations looking to use protected data for public health purposes.
To request an application pack or discuss a request for UKHSA data you would like to submit, contact.

## Acknowledgements

We thank the members of the 2023/24 measles incident UKHSA Data Epidemiology and Analytics cell for their insight and expertise that shaped this work and its usefulness. We thank UKHSA data operations colleagues for their work in the access, processing and maintenance of the data used in this paper.

