## Supplementary material for "Nowcasting cases and trends during the measles 2023/24 outbreak in England"

#### Section 1: Data quarantine

Some cases with a positive test may be set aside for review instead of being immediately added to the line list. Quarantined cases are flagged for review by criteria such as being a possible vaccine-associated case or having a positive local lab test but a negative reference lab test. These cases are then reviewed by clinicians and epidemiologists, sometimes needing a confirmatory reference lab test, and either discarded if judged to be a vaccine-associated case or a non-case, or added into the line list.

#### Section 2: Model hyperparameter and model structure tuning

For all models presented in the main paper, model hyperparameter and model structure tuning of the models presented in the main paper were determined by optimising the national daily weighted interval score (WIS) on a log scale. Log WIS was averaged over all predictions in the last four weeks, the nowcast horizon for which case count predictions were made, or eight weeks for the report date model which was not used for case count estimates, only trends. This included training length, maximum delay (if applicable) and choice of smooths. The smooths tested and their associated hyperparameters tuned were thin plate regression (number of basis functions), cubic regression (number of knots) and Gaussian process smooths (power of exponential power covariance function). All scores are provided as supplementary data.

Due to time constraints when developing the operational model, we did not explore alternative model structures thoroughly. Hence, for the retrospective model, we additionally test simpler delay structures to assess performance over the whole outbreak. The following model structures were considered:

Model structure 1: Disaggregating delay by both test type and region (as in the operational model)

$$\log\left(\mu_{t,d,\text{test type},\text{region}}\right) = \beta_0 + s_{1,\text{region}}(t) + s_{2,\text{test type}}(d) + s_{3,\text{test type},\text{region}}(d) \\ +\omega_1\left(\text{wday}(t)\right) + \omega_2\left(\text{wday}(t+d)\right) + \omega_3(\text{region})$$

Model structure 2: Disaggregating delay by test type only

$$\log\left(\mu_{t,d,\text{test type},\text{region}}\right) \\ = \beta_0 + s_{1,\text{region}}(t) + s_{2,\text{test type}}(d) + \omega_1\left(\text{wday}(t)\right) + \omega_2\left(\text{wday}(t+d)\right) \\ + \omega_3(\text{region})$$

Model structure 3: Disaggregating delay by region only

$$\log(\mu_{t,d,\text{region}}) = \beta_0 + s_{1,\text{region}}(t) + s_{2,\text{region}}(d) + \omega_1(\text{wday}(t)) + \omega_2(\text{wday}(t+d)) + \omega_3(\text{region})$$

Model structure 4: No disaggregation of delay

$$\log(\mu_{t,d,\text{region}}) = \beta_0 + s_{1,\text{region}}(t) + s_2(d) + \omega_1(\text{wday}(t)) + \omega_2(\text{wday}(t+d)) + \omega_3(\text{region})$$

Averaged over national daily scores, these model structures performed similarly in terms of log WIS, with the model with delay by region and test type slightly worse than the others (Supplementary Table 1). For London and the West Midlands, the model with delay by region outperforms other models, but the order of other models differs, with test type more important in London but not the West Midlands. For the analysis in this paper, we choose the best model and hyperparameters according to the national daily scoring.

### Section 3: Growth rates

#### Methods

Growth rates were not presented operationally during the outbreak, but we consider them here as an alternative output for assessing trends. We focus on the growth rate on the last day of prediction, representing the most recent trend.

$$r_{\text{region}}(T) = \left.\frac{dN_{t,\text{region}}}{dt}\right|_{t=T} \tag{1}$$

Regional growth rate samples are calculated from the fitted model using the *derivative_samples* function from the *gratia* package [15]. These derivative samples are computed via finite differences of posterior draws from the model's expected response values. National growth rate samples are estimated by the sum of the regional growth rate samples, weighted by the proportion of cases within each region. To account for noisy data, the proportion of cases per region is calculated as a rolling average of regional case counts divided by the rolling average of national case counts, using a 28-day window. Prediction intervals are extracted from the distribution of growth rate samples.

#### Results

Final day growth rate estimates are mostly positive during the sharp incline of the England and West Midlands epidemic curves, and negative or near 0 during the slow decline in the West Midlands (Supplementary Figure 29). However, during less well-defined growth and decline phases, estimates often have wide prediction intervals that overlap with zero. This is less so for the report date model, which has narrower prediction intervals, particularly in London, where it is less responsive to local peaks and predicts a positive growth rate for most of the London epidemic until it begins to decline. Other models are more sensitive to local trend changes, but as a result have wider prediction intervals. The report date model also exhibits a delayed response to prominent trend changes such as at the West Midlands peak, where its transition from a confident positive growth rate is lagged by one week compared to other models.

#### Supplementary data

Scores for the tuning of hyperparameters and structures for the retrospective, baseline and report date models are given as a csv, with log WIS values. For rows relating to the retrospective model, the column “retrospective_model_structure” refers to the model structures in Supplementary Section 2. The columns “s1_smooth_type” and “s1_basis_size” refer to the $s_1$ smooths in the model formulae, and if “s1_smooth_type” is a Gaussian process, “s1_gp_covariance_power” is the power of the exponential power covariance function. If applicable, “s2_basis_size” refer to the cubic regression $s_2$ smooths in the model formulae.

#### Code

Code for the models and results in the paper is available at https://github.com/maria-tang/measles-nowcast.

#### Supplementary plots

| Geography | Disaggregation of delay in model | Smooth type | Log WIS |
|---|---|---|---|
| England | **Region** | **tp** | **0.115** |
| | None | gp | 0.116 |
| | Test type | gp | 0.116 |
| | Region and test type | gp | 0.122 |
| London | **Region** | **tp** | **0.615** |
| | Region and test type | cr | 0.644 |
| | Test type | gp | 0.746 |
| | None | gp | 0.755 |
| West Midlands | **Region** | **gp** | **0.980** |
| | None | gp | 0.995 |
| | Test type | gp | 0.995 |
| | Region and test type | cr | 1.060 |

Supplementary Table 1 (referenced in Supplementary Section 2): Retrospective model scores for different model structures, smooth types and geography.

| Report dates available | % of cases |
|---|---|
| Local lab only | 18.9 |
| Reference lab only | 23.2 |
| Local & reference lab | 54.3 |
| Neither | 3.5 |

Supplementary Table 2: Percentage of cases in the line list with one, both or neither of a local lab and reference lab test date recorded.

| Type of date | Minimum | Maximum | Non-missing count | Missing count | Missing (%) | Days since symptom onset median (90% quantile range) | Days since symptom onset < 0 (%) |
|---|---|---|---|---|---|---|---|
| Symptom onset date | 2023-10-01 | 2024-09-08 | 2734 | 0 | 0.0 | - | - |
| Local lab received date | 2023-10-03 | 2024-09-21 | 2002 | 732 | 26.8 | 6 [2, 20] | 0.8 |
| Reference lab report date | 2023-10-23 | 2024-10-05 | 2120 | 614 | 22.5 | 26 [10, 69] | 0.1 |
| Earliest report date | 2023-10-03 | 2024-10-05 | 2637 | 97 | 3.5 | 8 [2, 47] | 0.7 |

Supplementary Table 3: Summary statistics for the date fields in the measles line list, based on data extracted on 8 October 2024 and filtered to include cases with symptom onset date up to 8 September 2024. Data before this maximum symptom onset date is assumed to be mostly complete.

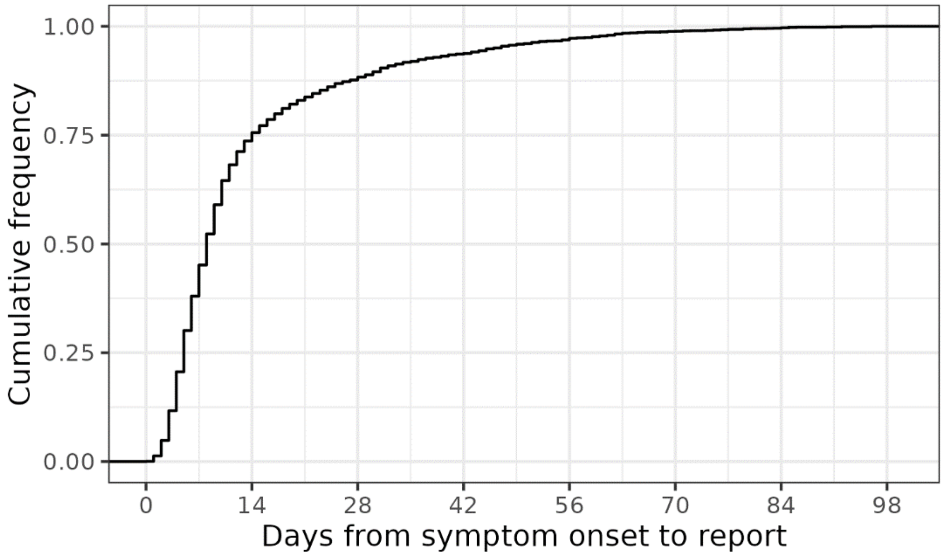


Supplementary Figure 1: Cumulative frequency of delays between earliest report and symptom onset date for all cases. A few cases have negative delays, which may indicate data quality issues. The minimum delay is -30 and the maximum is 172 days delay.

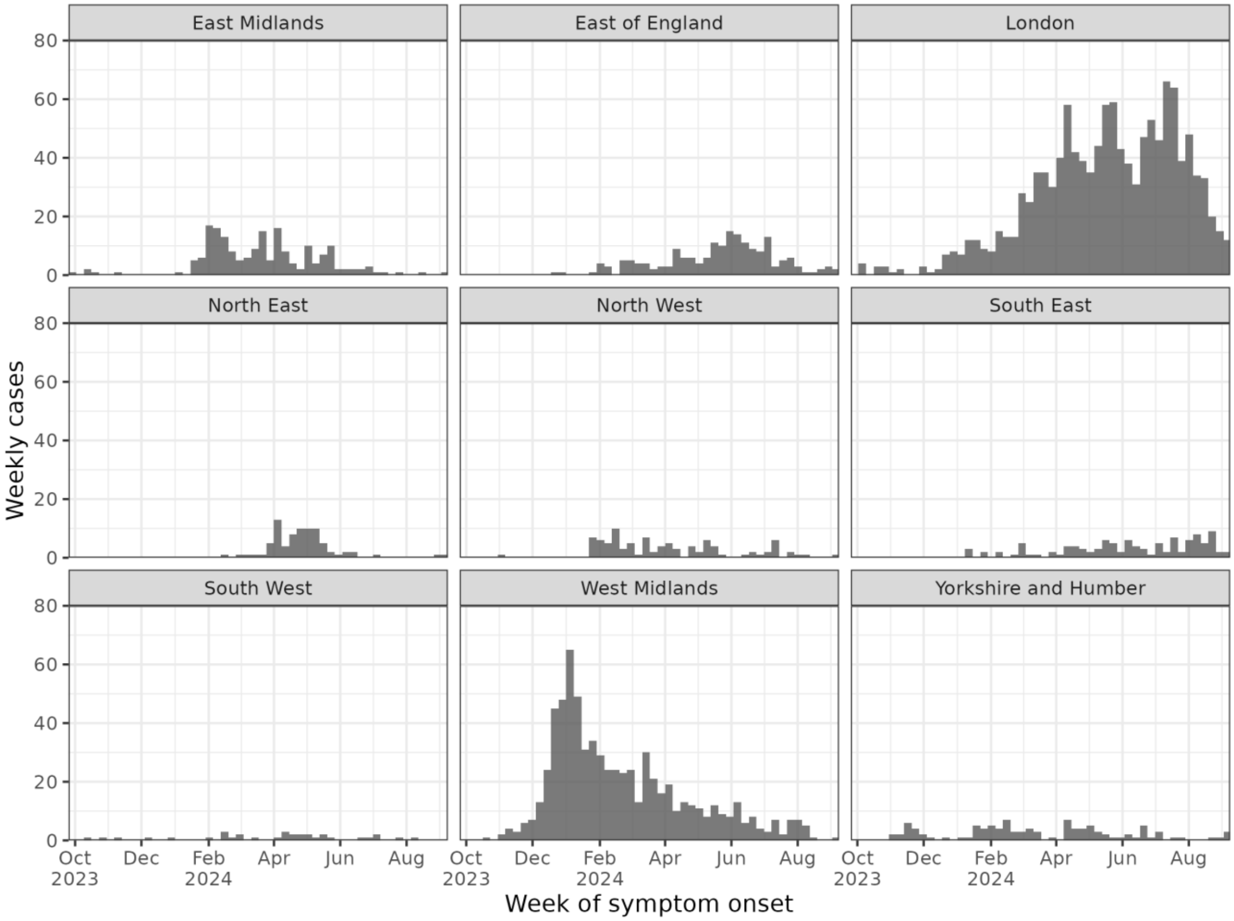


Supplementary Figure 2: Count of confirmed measles cases by region and week according to symptom onset date.

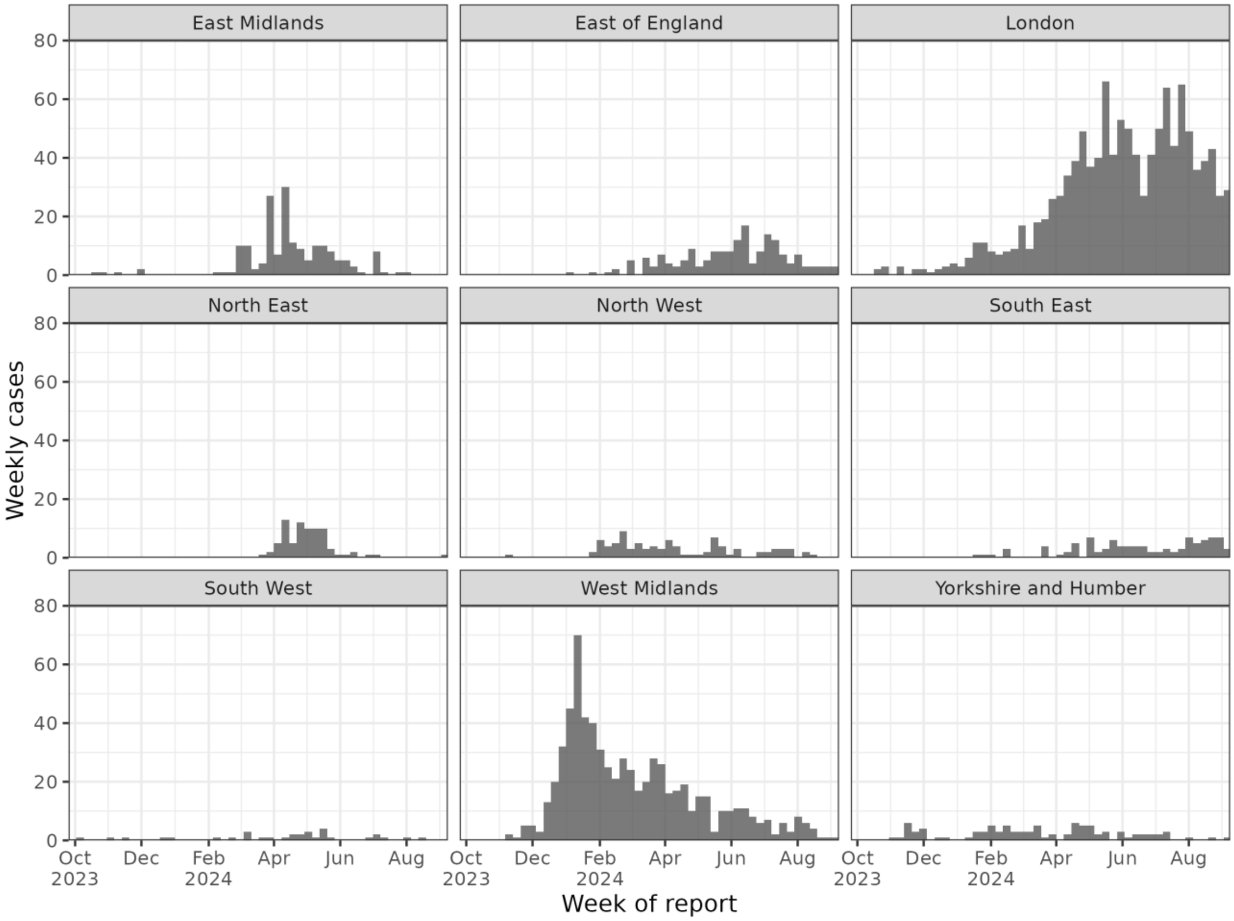


Supplementary Figure 3: Count of confirmed measles cases by region and week according to earliest report date.

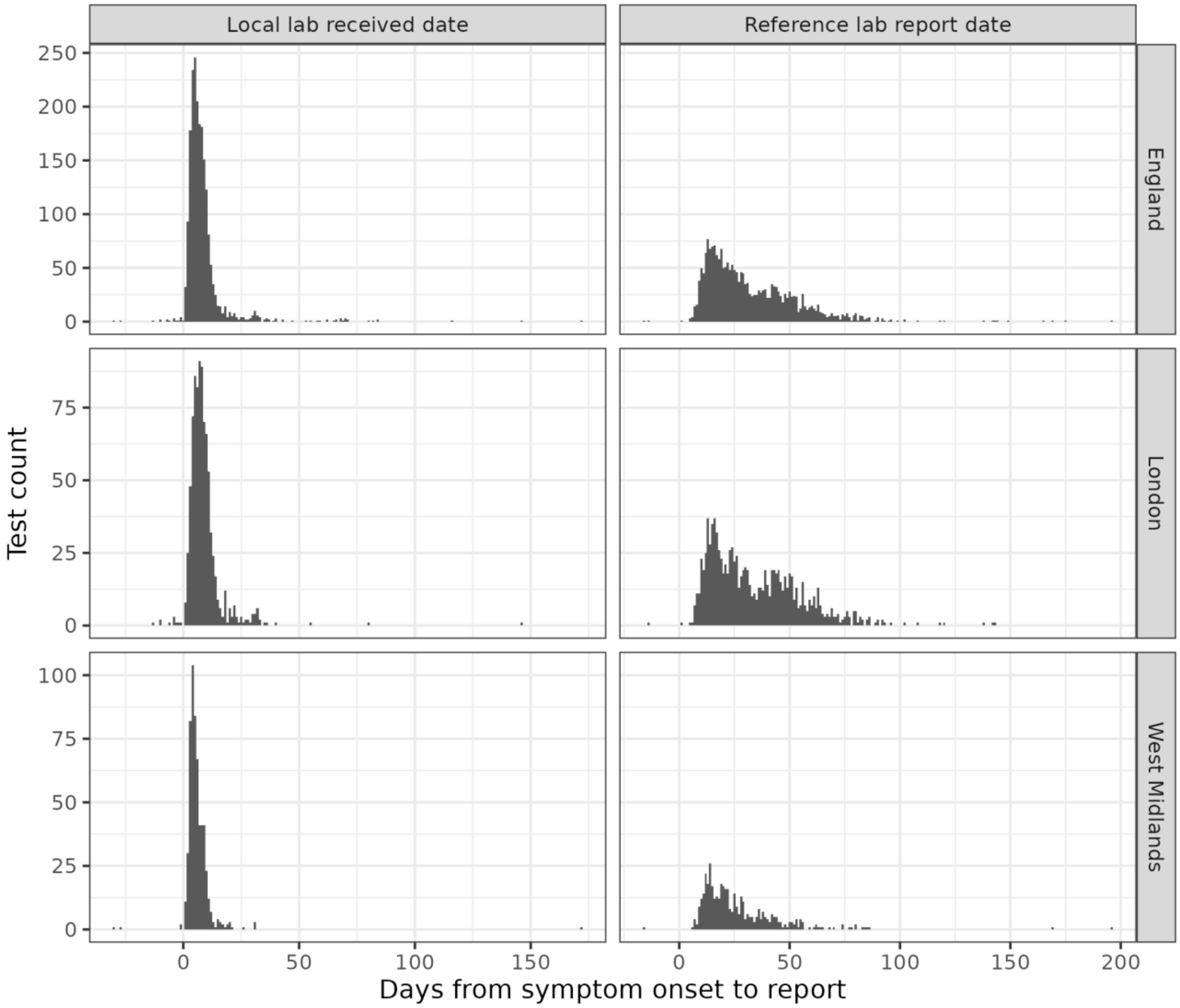


Supplementary Figure 4: Distribution of test reporting delays by geographic region and test type. This includes all tests, including multiple tests from the same confirmed case. A few cases have negative delays, which may indicate data quality issues.

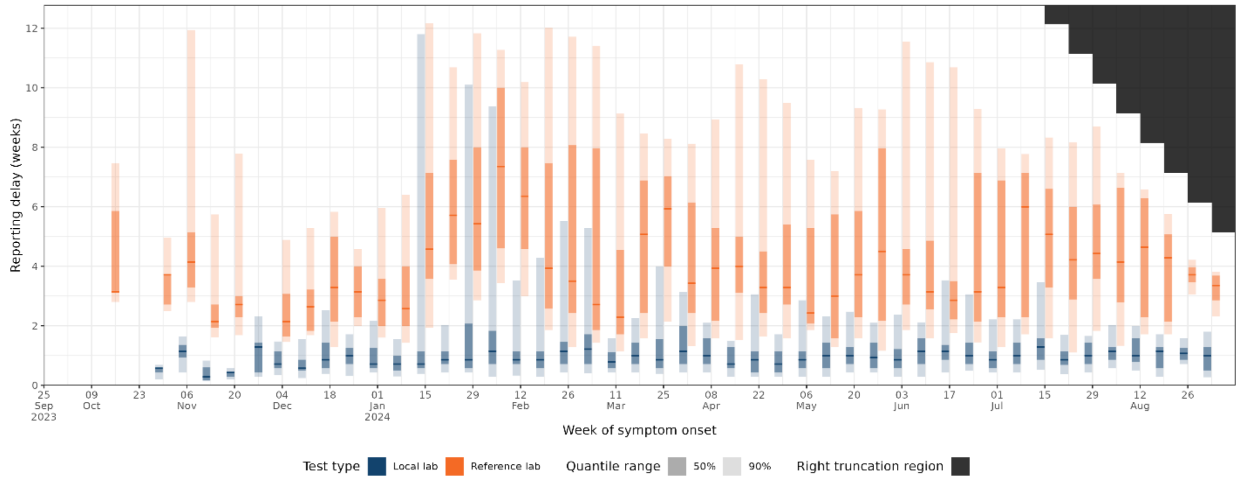


Supplementary Figure 5: Reporting delay for cases by test type and week, defined as the difference between symptom onset and earliest report date. Negative delays are excluded. Small count subgroups with $\leq 5$ cases in a week are excluded. The right truncation region shows the values that cannot be observed due to right truncation of the data.

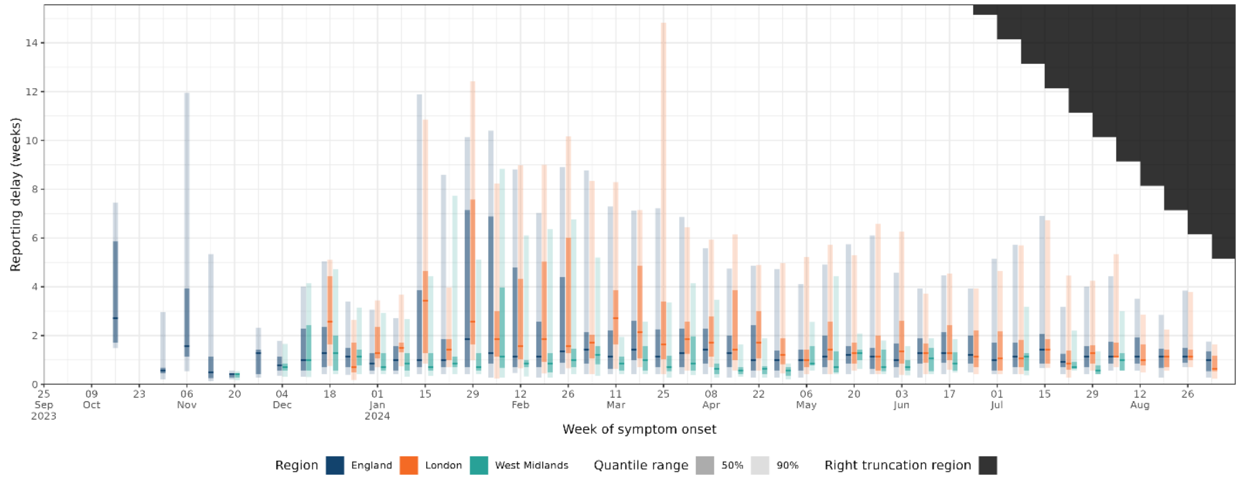


Supplementary Figure 6: Reporting delay for cases by region and week, defined as the difference between symptom onset and earliest report date. Negative delays are excluded. Small count subgroups with $\leq 5$ cases in a week are excluded. The right truncation region shows the values that cannot be observed due to right truncation of the data.

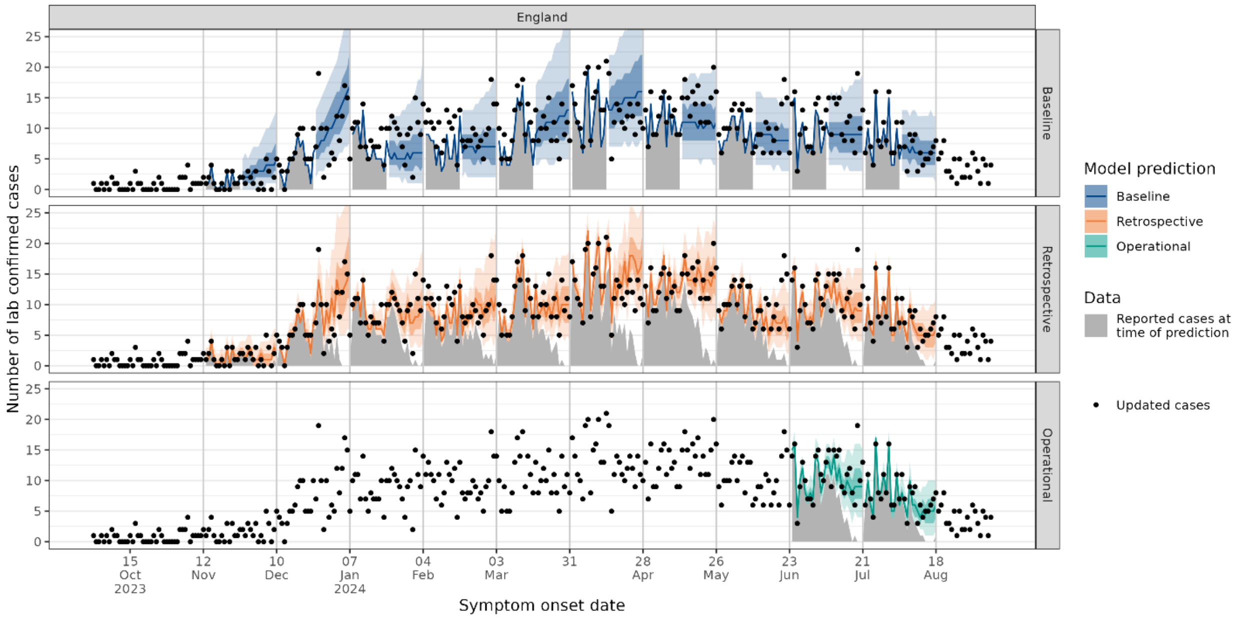


Supplementary Figure 7: Daily national predictions (median, 50% and 90% prediction intervals) for models re-fitted on new available data each week. Every fourth model fit is shown for clarity, with the first one fitted on data up to 10 December 2023. The operational model is shown only for the period it was used during the outbreak.

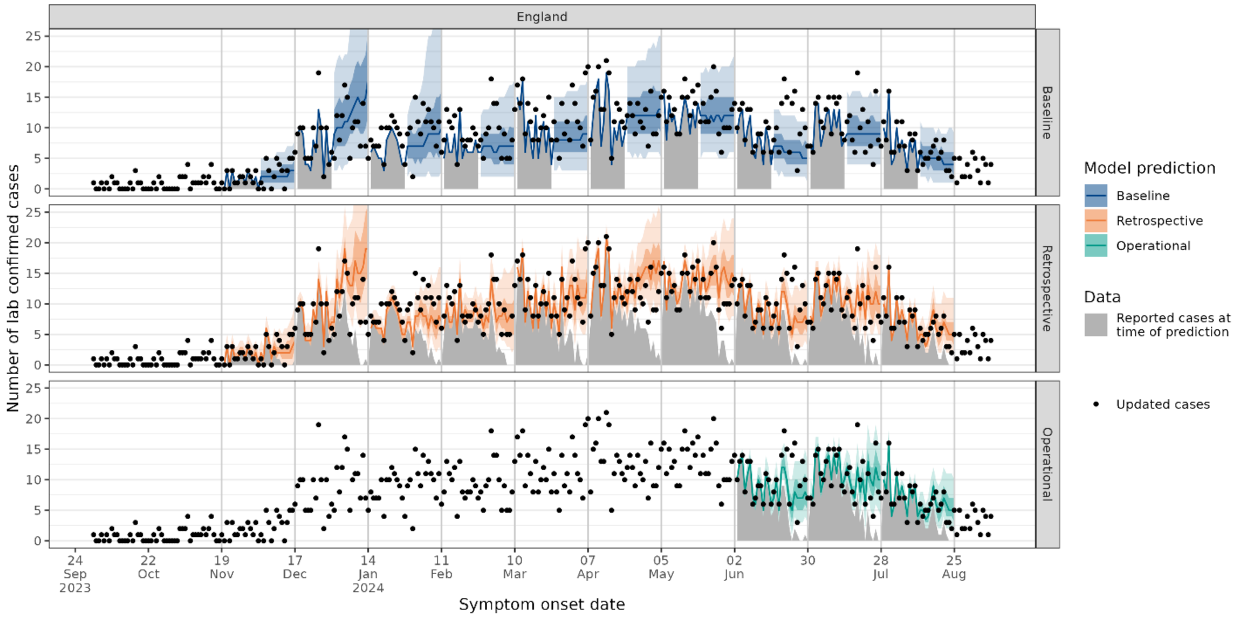


Supplementary Figure 8: Daily national predictions (median, 50% and 90% prediction intervals) for models re-fitted on new available data each week. Every fourth model fit is shown for clarity, with the first one fitted on data up to 17 December 2023. The operational model is shown only for the period it was used during the outbreak.

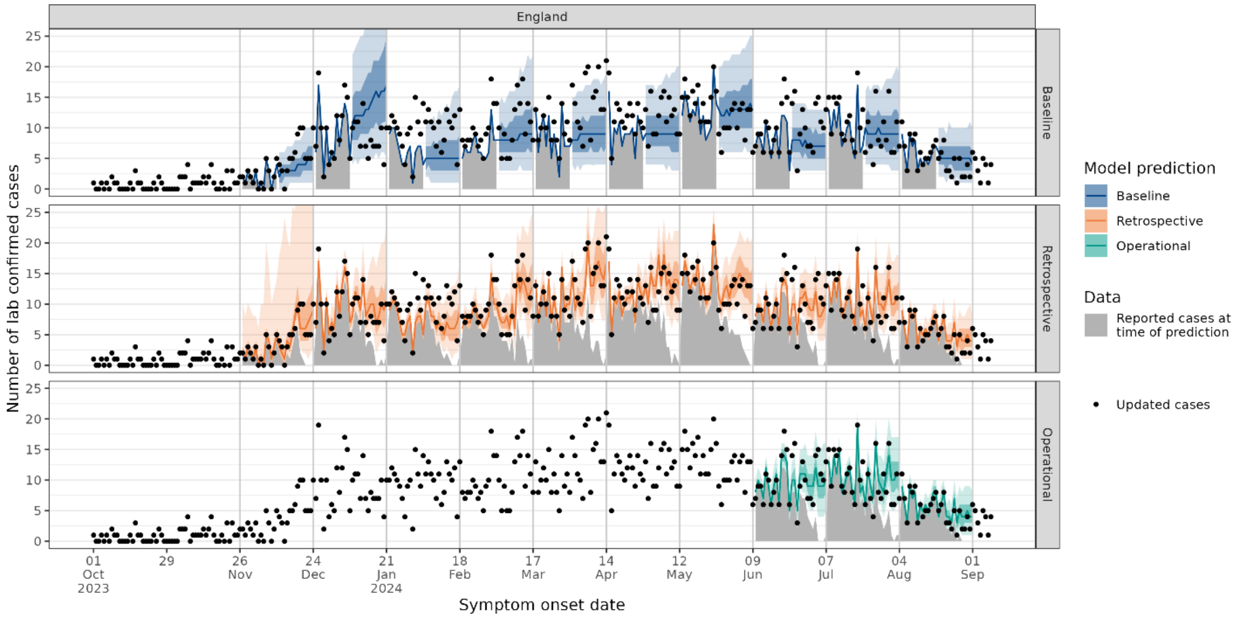


Supplementary Figure 9: Daily national predictions (median, 50% and 90% prediction intervals) for models re-fitted on new available data each week. Every fourth model fit is shown for clarity, with the first one fitted on data up to 24 December 2023. The operational model is shown only for the period it was used during the outbreak.

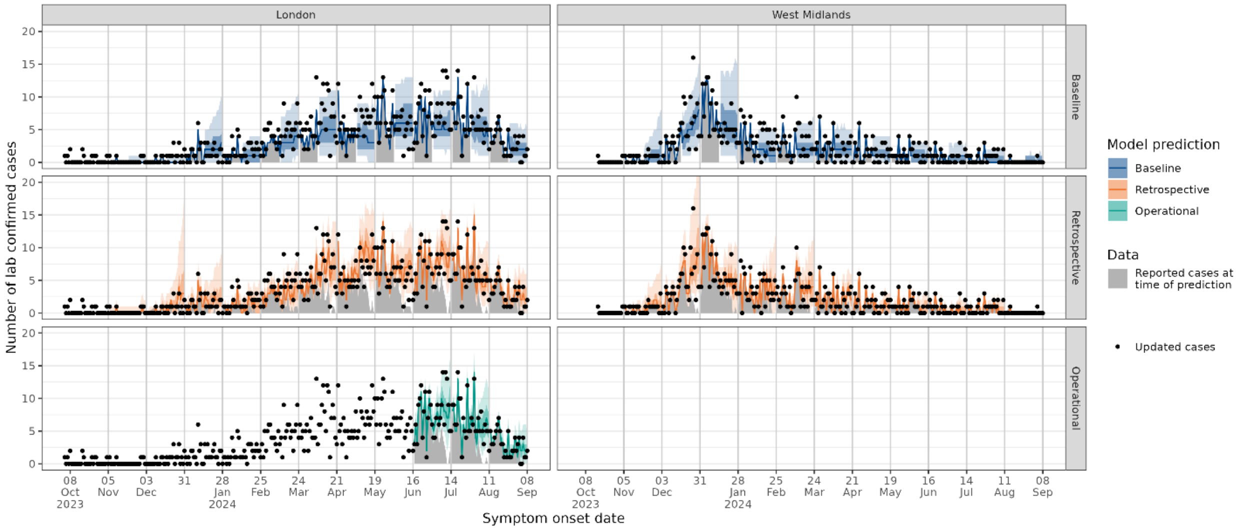


Supplementary Figure 10: Daily predictions (median, 50% and 90% prediction intervals) in London and the West Midlands for models re-fitted on new available data each week. Every fourth model fit is shown for clarity, with the first one fitted on data up to 3 December 2023. Predictions for all other weeks during the study period are given in Supplementary Figure 11 to Supplementary Figure 13. The operational model is shown only for the period it was used during the outbreak.

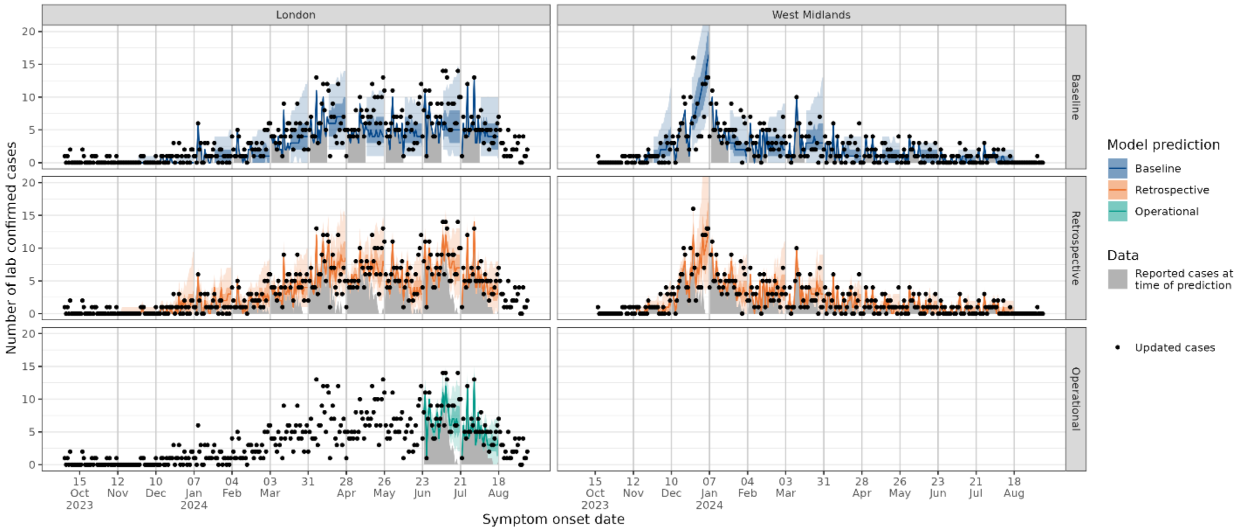


Supplementary Figure 11: Daily predictions (median, 50% and 90% prediction intervals) in London and the West Midlands for models re-fitted on new available data each week. Every fourth model fit is shown for clarity, with the first one fitted on data up to 10 December 2023. The operational model is shown only for the period it was used during the outbreak.

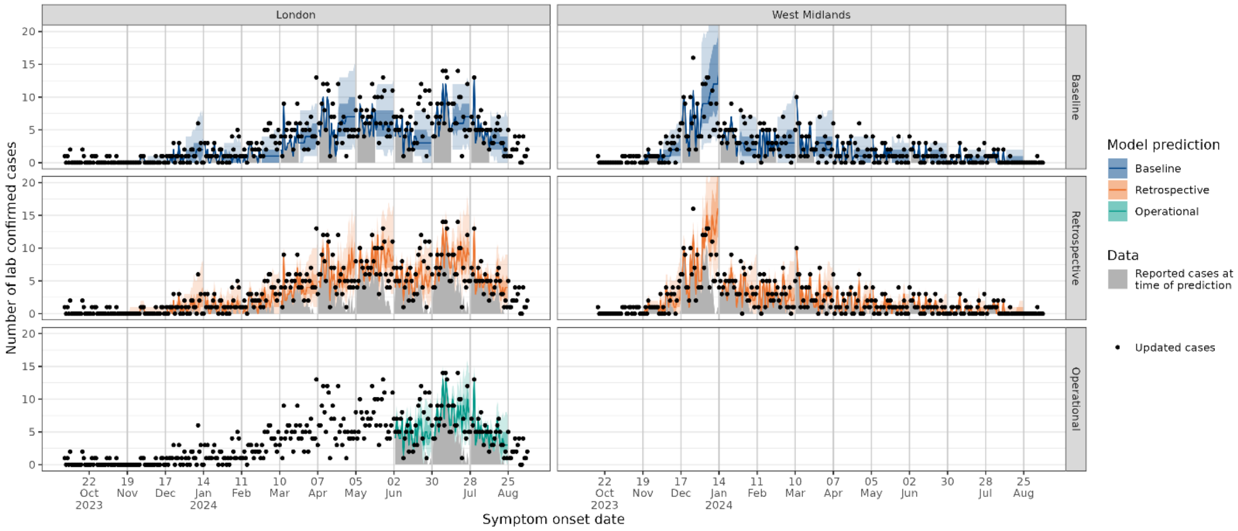


Supplementary Figure 12: Daily predictions (median, 50% and 90% prediction intervals) in London and the West Midlands for models re-fitted on new available data each week. Every fourth model fit is shown for clarity, with the first one fitted on data up to 17 December 2023. The operational model is shown only for the period it was used during the outbreak.

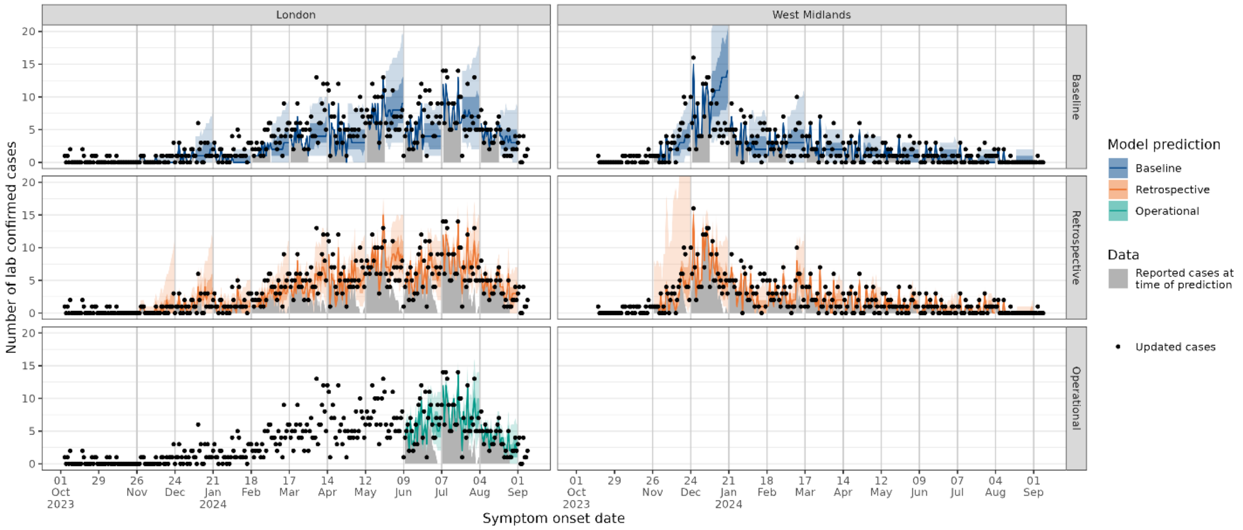


Supplementary Figure 13: Daily predictions (median, 50% and 90% prediction intervals) in London and the West Midlands for models re-fitted on new available data each week. Every fourth model fit is shown for clarity, with the first one fitted on data up to 24 December 2023. The operational model is shown only for the period it was used during the outbreak.

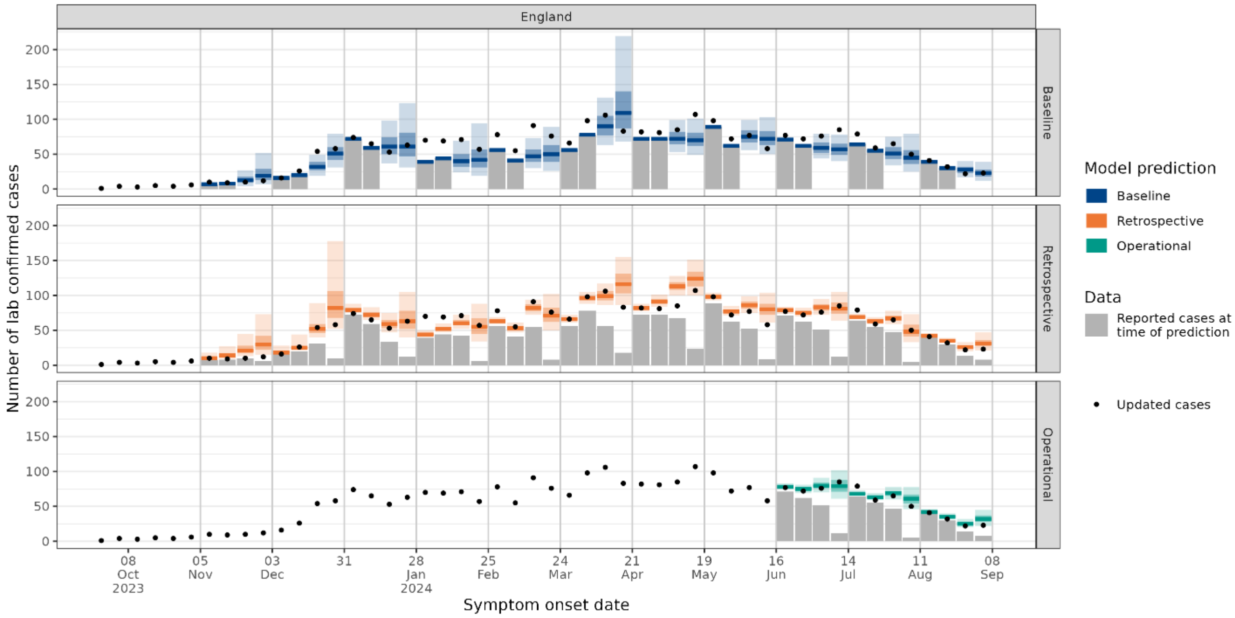


Supplementary Figure 14: Weekly national predictions (median, 50% and 90% prediction intervals) for models re-fitted on new available data each week. Every fourth model fit is shown for clarity, with the first one fitted on data up to 3 December 2023. Predictions for all other weeks during the study period are given in Supplementary Figure 15 to Supplementary Figure 17. The operational model is shown only for the period it was used during the outbreak.

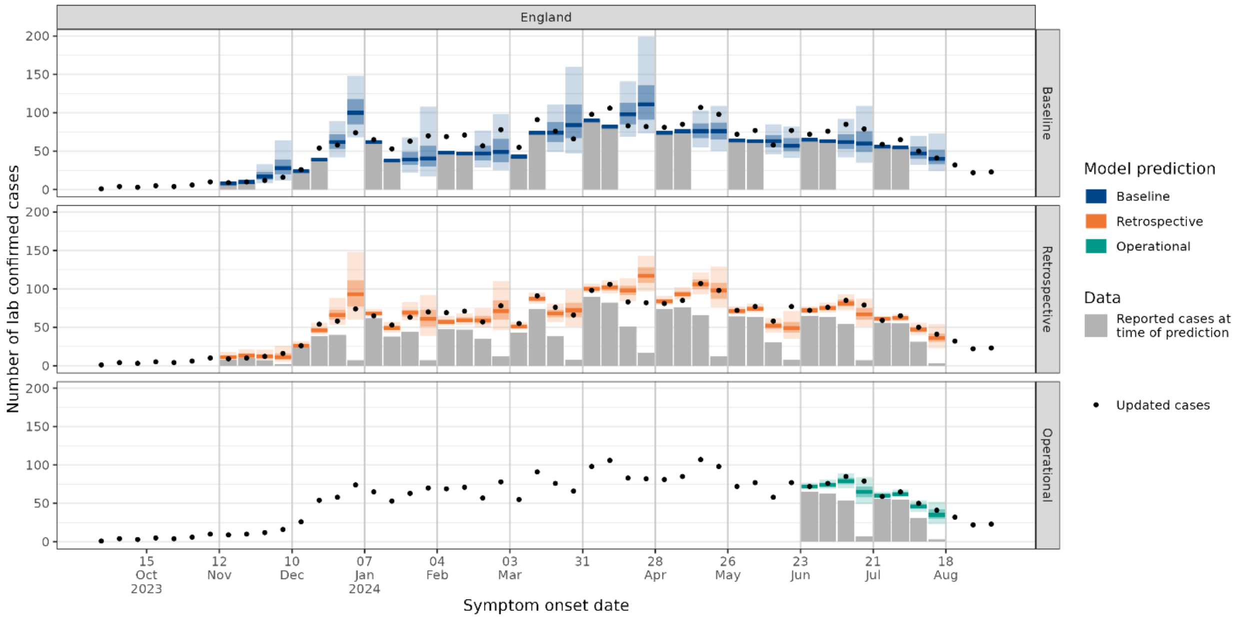


Supplementary Figure 15: Weekly national predictions (median, 50% and 90% prediction intervals) for models re-fitted on new available data each week. Every fourth model fit is shown for clarity, with the first one fitted on data up to 10 December 2023. The operational model is shown only for the period it was used during the outbreak.

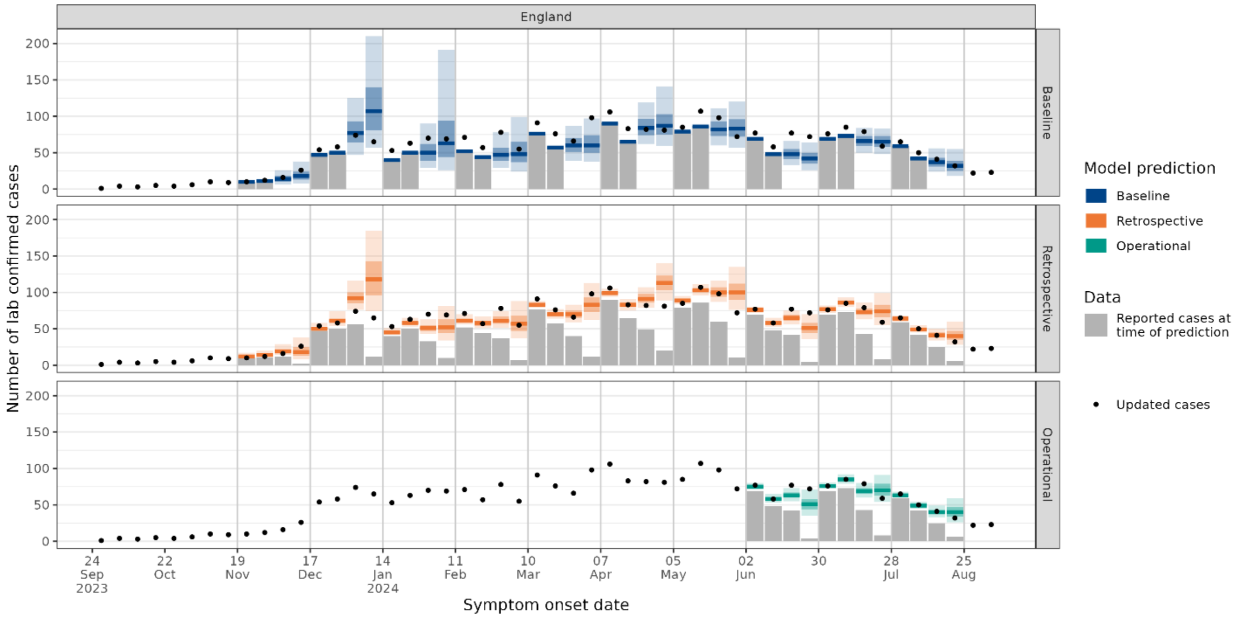


Supplementary Figure 16: Weekly national predictions (median, 50% and 90% prediction intervals) for models re-fitted on new available data each week. Every fourth model fit is shown for clarity, with the first one fitted on data up to 17 December 2023. The operational model is shown only for the period it was used during the outbreak.

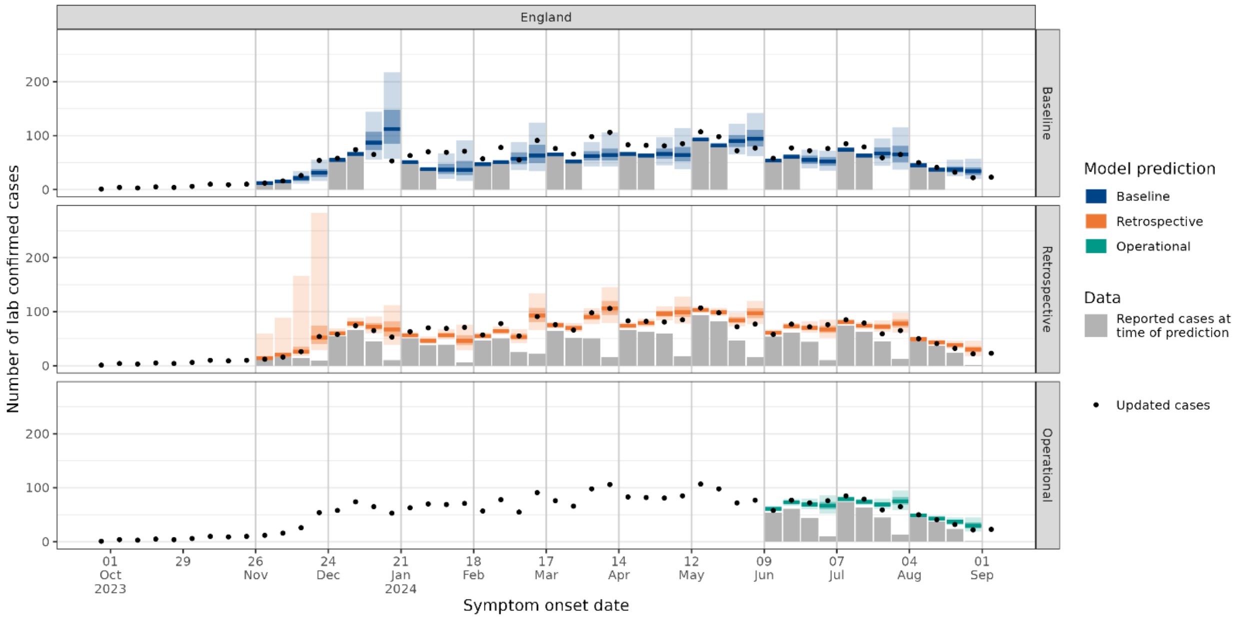


Supplementary Figure 17: Weekly national predictions (median, 50% and 90% prediction intervals) for models re-fitted on new available data each week. Every fourth model fit is shown for clarity, with the first one fitted on data up to 24 December 2023. The operational model is shown only for the period it was used during the outbreak.

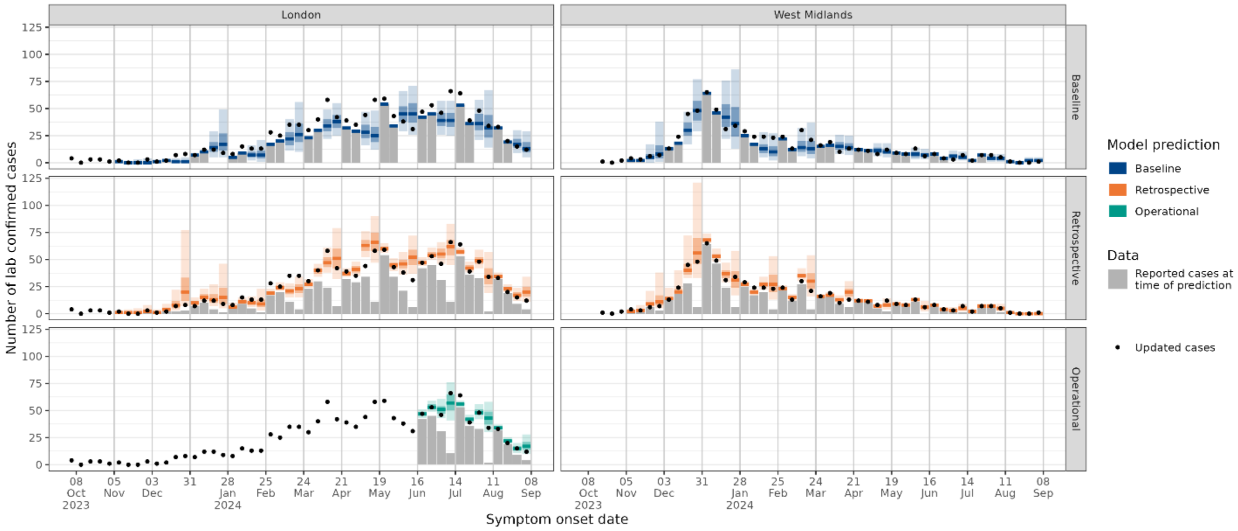


Supplementary Figure 18: Weekly predictions (median, 50% and 90% prediction intervals) in London and the West Midlands for models re-fitted on new available data each week. Every fourth model fit is shown for clarity, with the first one fitted on data up to 3 December 2023. Predictions for all other weeks during the study period are given in Supplementary Figure 19 to Supplementary Figure 21. The operational model is shown only for the period it was used during the outbreak.

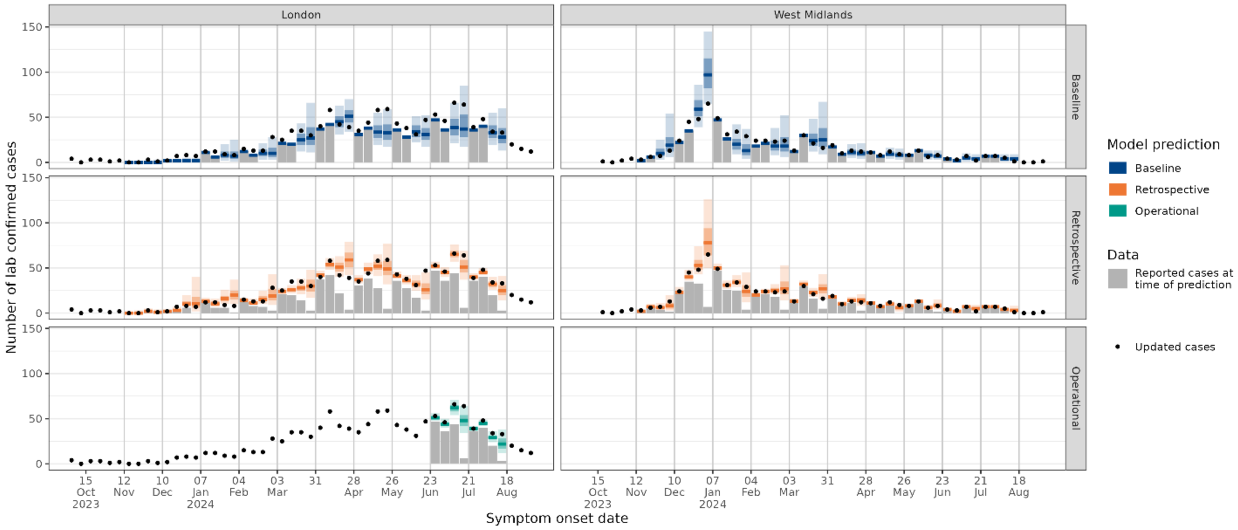


Supplementary Figure 19: Weekly predictions (median, 50% and 90% prediction intervals) in London and the West Midlands for models re-fitted on new available data each week. Every fourth model fit is shown for clarity, with the first one fitted on data up to 10 December 2023. The operational model is shown only for the period it was used during the outbreak.

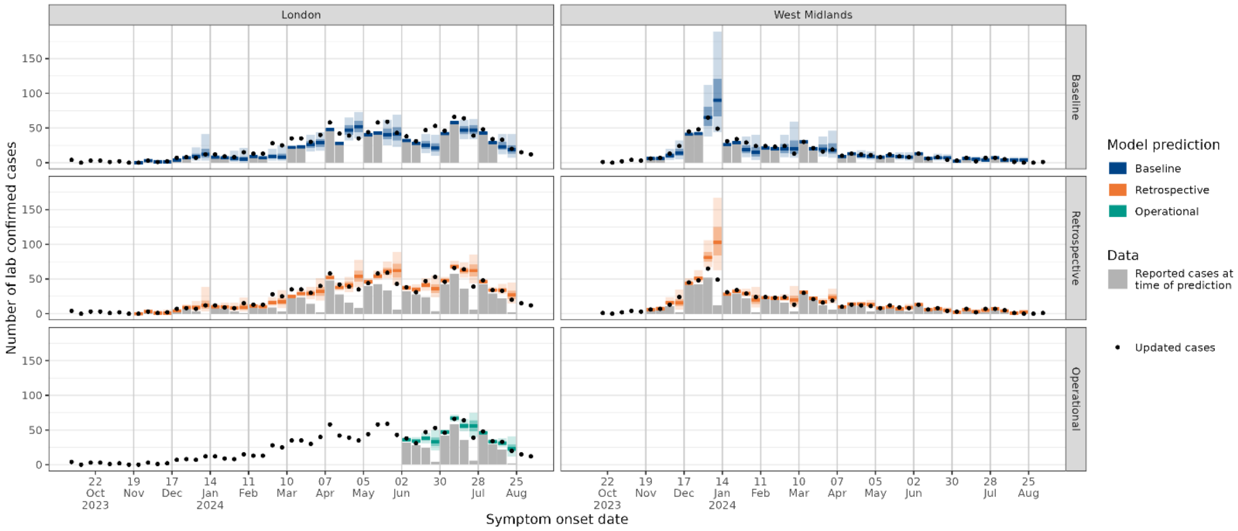


Supplementary Figure 20: Weekly predictions (median, 50% and 90% prediction intervals) in London and the West Midlands for models re-fitted on new available data each week. Every fourth model fit is shown for clarity, with the first one fitted on data up to 17 December 2023. The operational model is shown only for the period it was used during the outbreak.

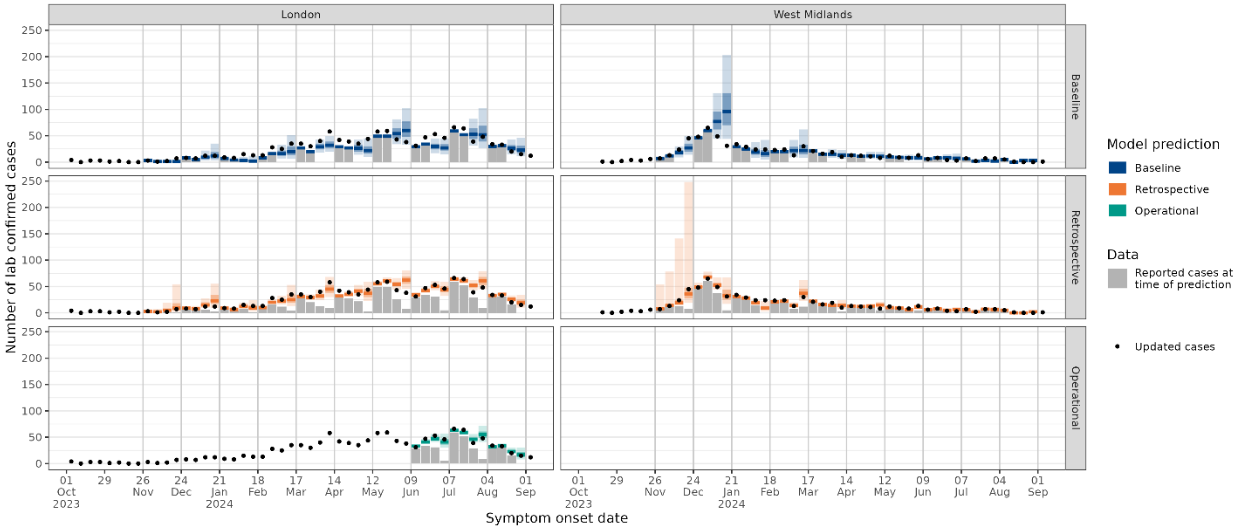


Supplementary Figure 21: Weekly predictions (median, 50% and 90% prediction intervals) in London and the West Midlands for models re-fitted on new available data each week. Every fourth model fit is shown for clarity, with the first one fitted on data up to 24 December 2023. The operational model is shown only for the period it was used during the outbreak.

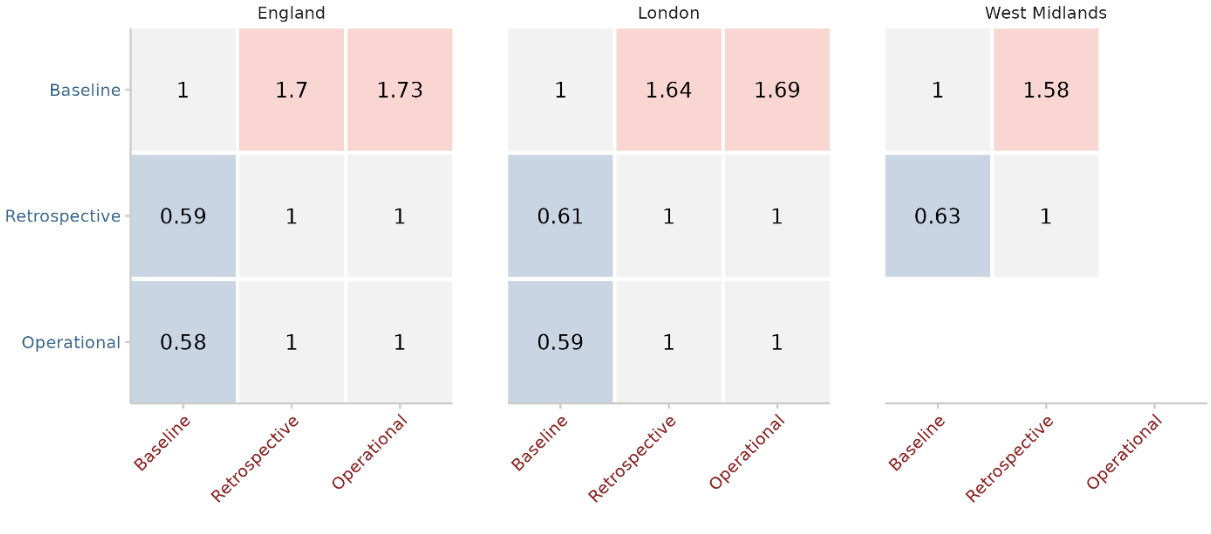


Supplementary Figure 22: Pairwise comparison of daily model scores for England, London, and the West Midlands, using ratio of mean daily log weighted interval scores. Values below 1 in blue represent the model in blue along the row performing better than the model in red along the column, and vice versa for values above 1 in red. Darker shaded squares represent a greater difference between models. The comparison between the operational model and other models is only for the weeks when the operational model was used.

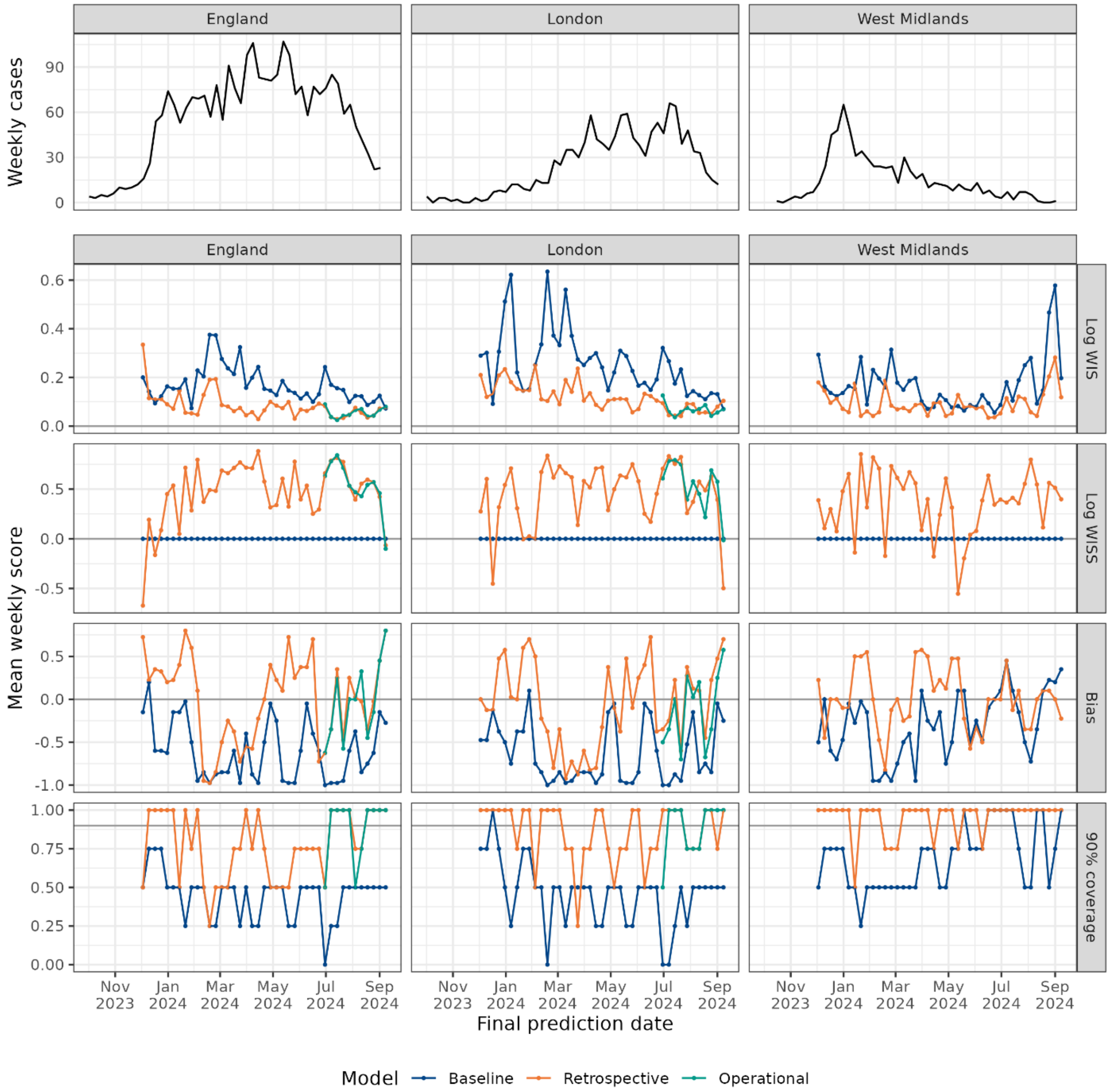


Supplementary Figure 23: Weekly cases over time in England, London and the West Midlands (top pane), and mean weekly scores by prediction end date and model: log weighted interval score (WIS), log weighted interval skill score (WISS), bias and 90% coverage. The comparison between the operational model and other models is only for the weeks when the operational model was used.

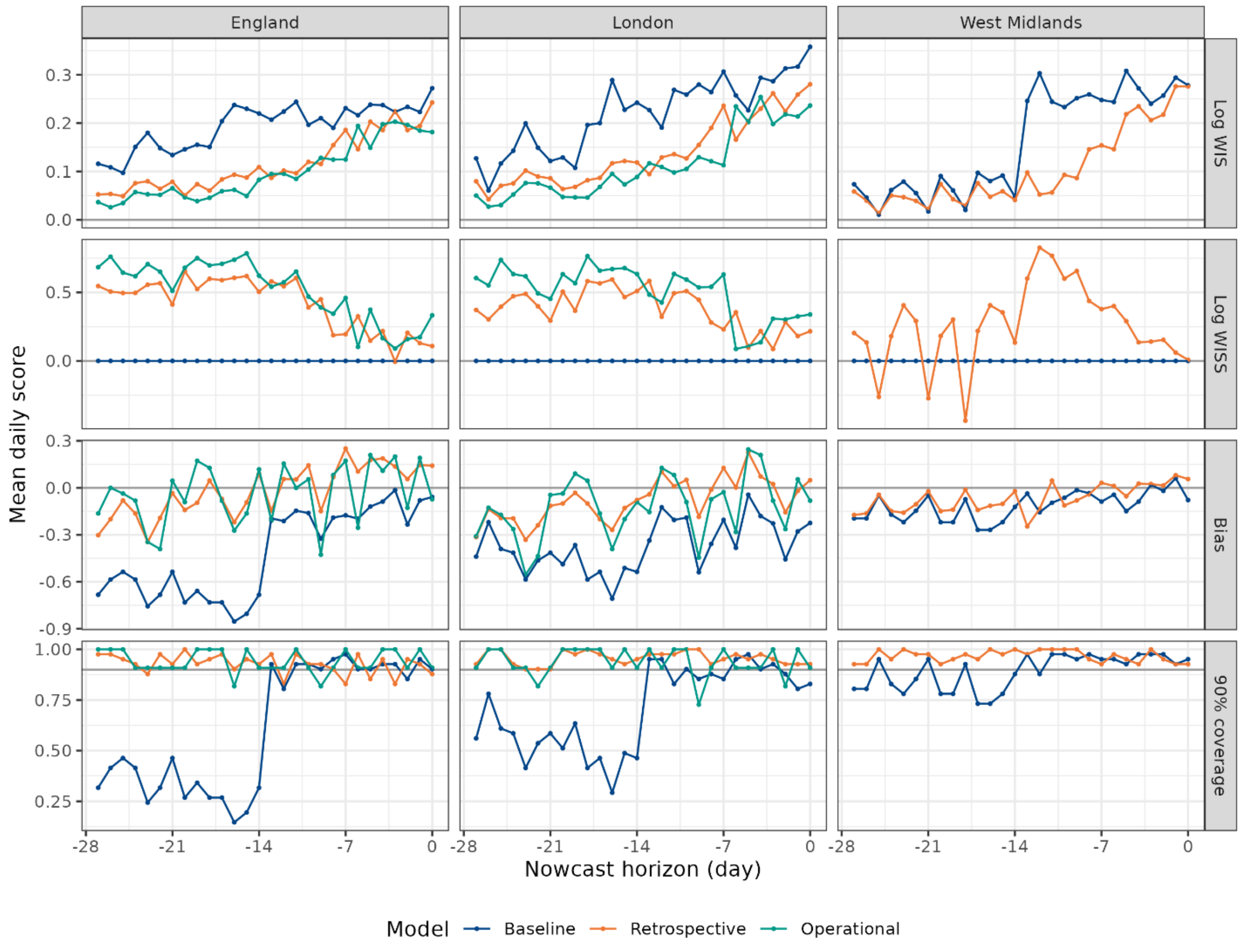


Supplementary Figure 24: Mean daily scores by day of nowcast horizon, the period over which predictions are made, where day 28 is the most recent day from which data is available. The panes show log weighted interval score (WIS), log weighted interval skill score (WISS), bias and 90% coverage. The comparison between the operational model and other models is only for the weeks when the operational model was used.

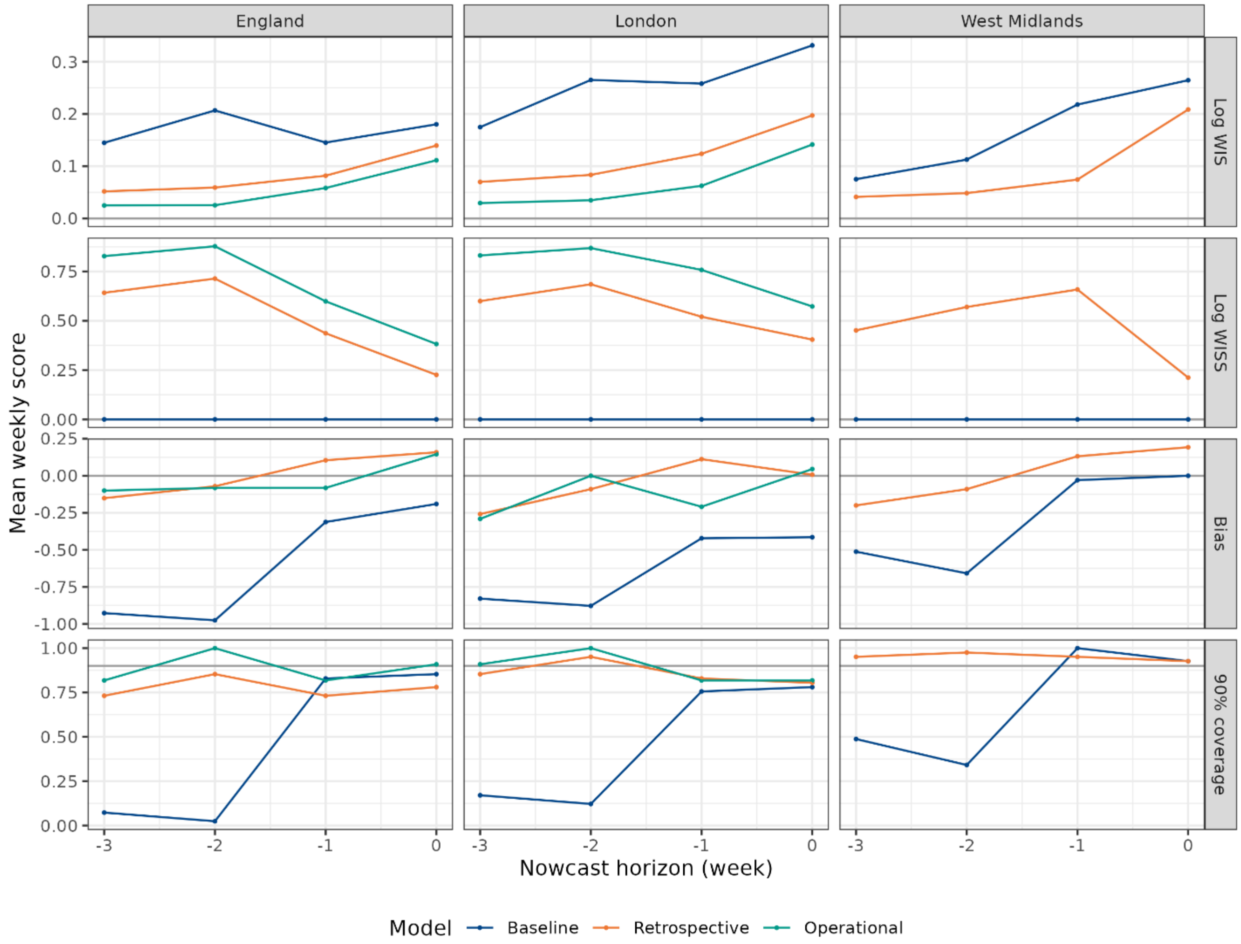


Supplementary Figure 25: Mean weekly scores by week of nowcast horizon, the period over which predictions are made, where week 4 is the most recent week from which data is available. The panes show log weighted interval score (WIS), log weighted interval skill score (WISS), bias and 90% coverage. The comparison between the operational model and other models is only for the weeks when the operational model was used.

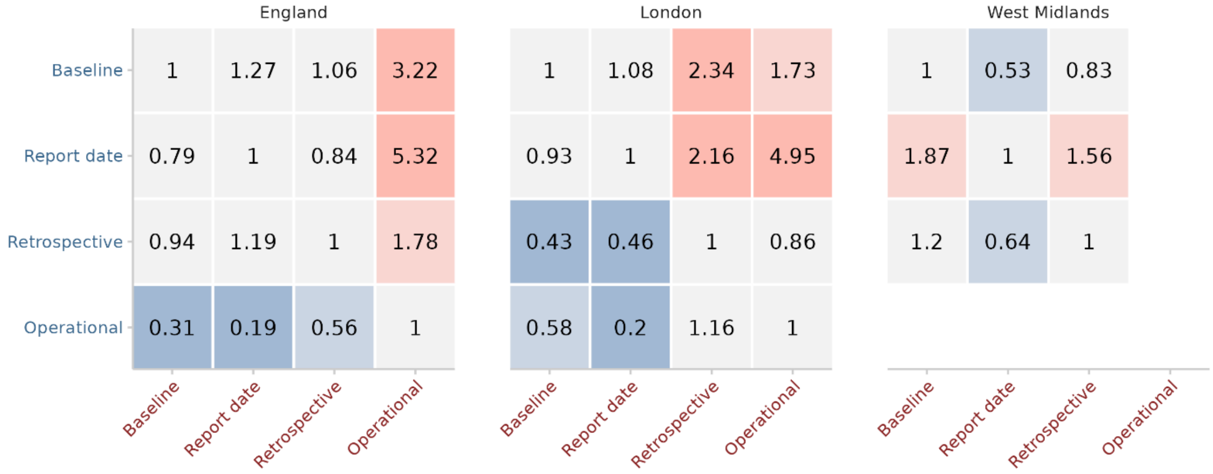


Supplementary Figure 26: Pairwise comparison of four-week trend categorisation scores for England, London, and the West Midlands, using ratio of mean ranked probability score (RPS). Values below 1 in blue represent the model in blue along the row performing better than the model in red along the column, and vice versa for values above 1 in red. Darker shaded squares represent a greater difference between models. Grey squares represent similar model scores, with ratio between 0.75 and 1.25. The comparison between the operational model and other models is only for the weeks when the operational model was used.

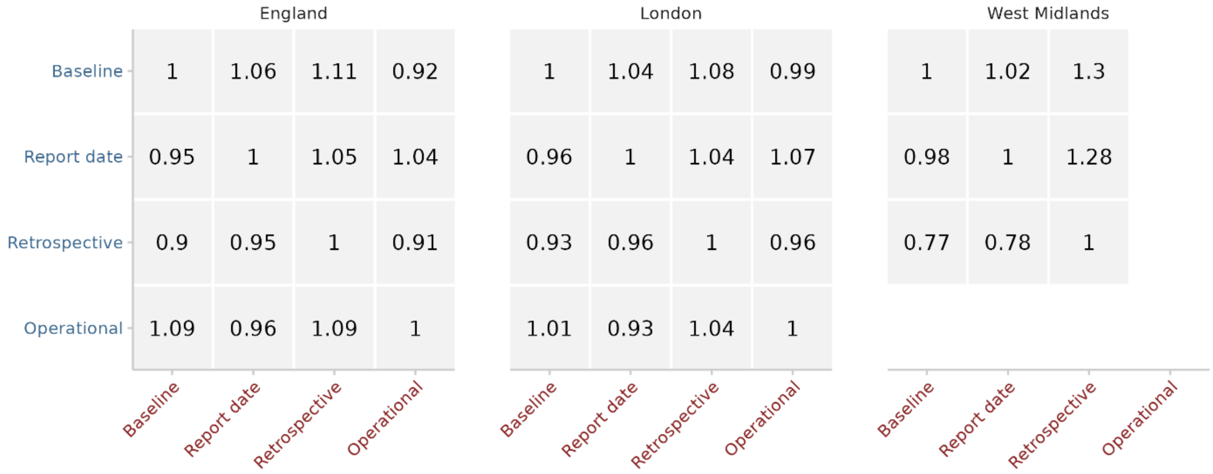


Supplementary Figure 27: Pairwise comparison of one-week trend categorisation scores for England, London, and the West Midlands, using ratio of mean ranked probability score (RPS). Values below 1 represent the model in blue along the row performing better than the model in red along the column, and vice versa for values above 1. The comparison between the operational model and other models is only for the weeks when the operational model was used.

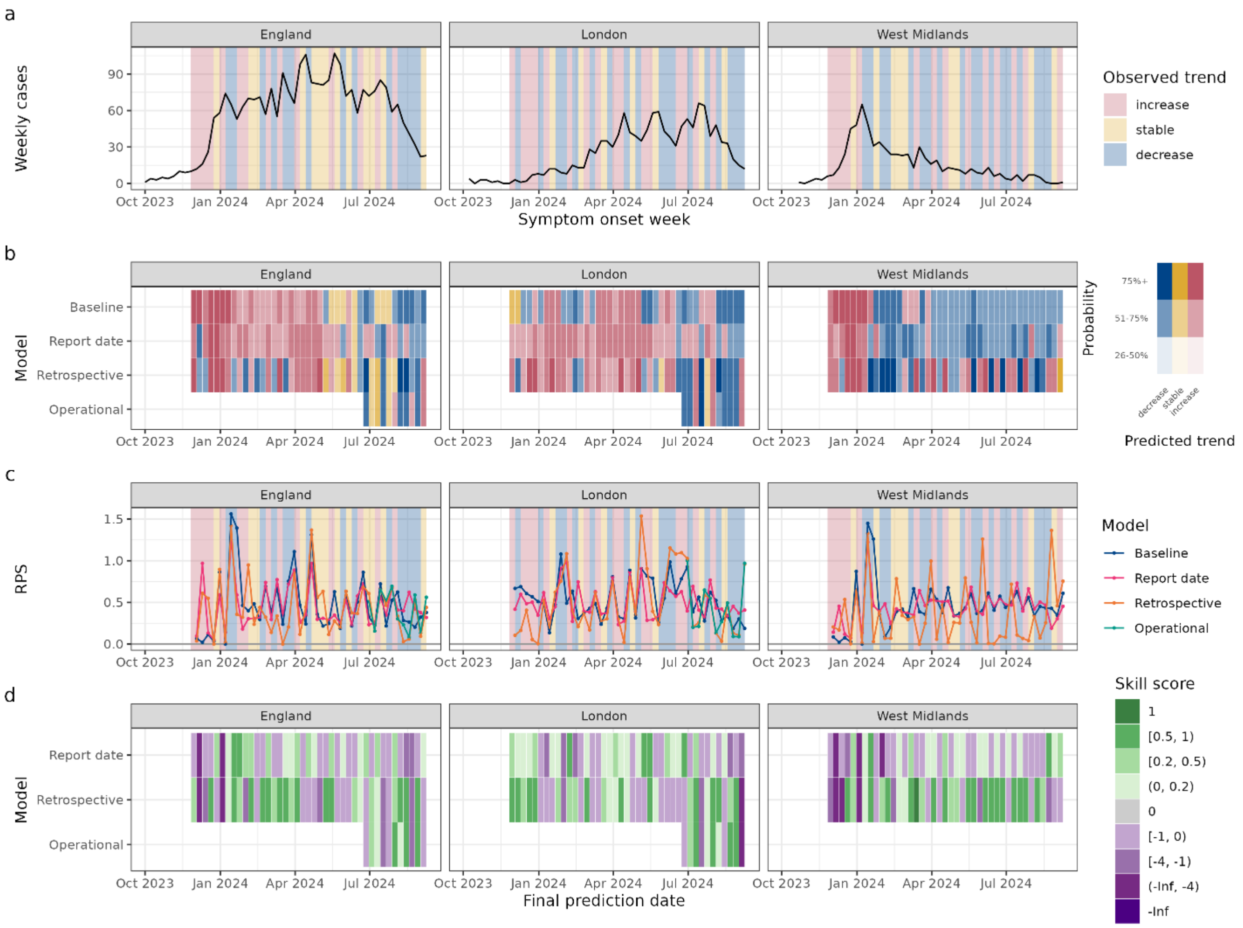


Supplementary Figure 28: (a) Weekly cases. (b) Predicted most likely one-week trend. (c) Ranked probability score (RPS) – lower is better. (d) Ranked probability skill score: green (better than baseline), grey (same), purple (worse). The operational model is shown only for the period it was used during the outbreak.

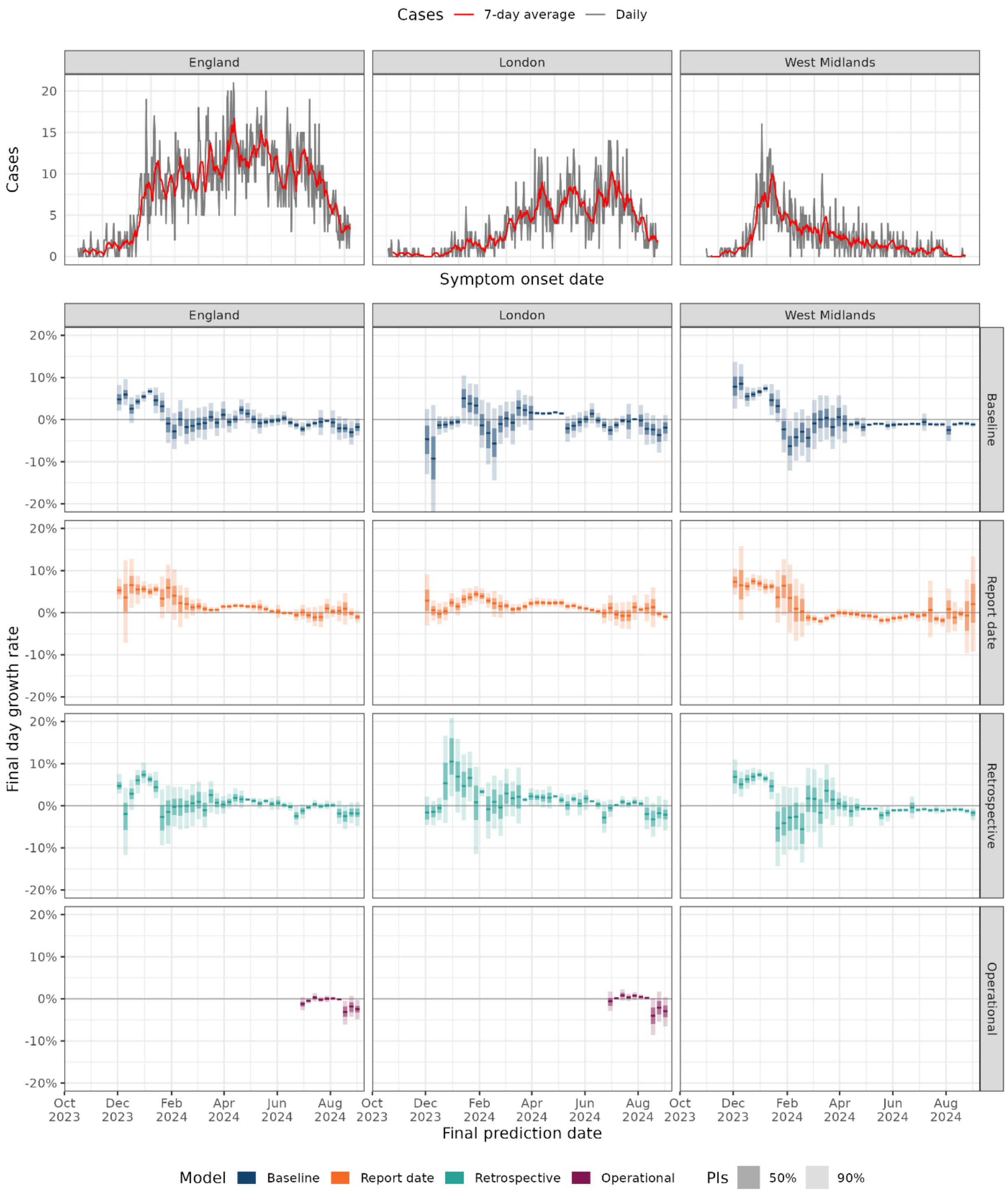


Supplementary Figure 29 (referenced in Supplementary Section 3): Daily cases over time with 7-day rolling average in England, London and the West Midlands (top pane), and growth rate estimates (median, 50% and 90% prediction intervals) for the final day of prediction by week and model. The operational model is shown only for the period it was used during the outbreak.
